# The impact of the COVID-19 pandemic on diagnoses of common mental health disorders in adults in Catalonia, Spain

**DOI:** 10.1101/2021.08.06.21261709

**Authors:** Berta Raventós, Andrea Pistillo, Carlen Reyes, Sergio Fernández-Bertolín, María Aragón, Anna Berenguera, Constanza Jacques-Aviñó, Laura Medina-Perucha, Edward Burn, Talita Duarte-Salles

**Author notes:** Joint senior authorship (Edward Burn, Talita Duarte-Salles). **Corresponding author** Talita Duarte-Salles, Fundació Institut Universitari per a la recerca a l’Atenció Primària de Salut Jordi Gol i Gurina (IDIAPJGol), Gran Via Corts Catalanes, 587 àtic, 08007 Barcelona - Spain.

## Abstract

**Objectives:** To investigate how incidence trends of anxiety and depressive disorders have been affected by the COVID-19 pandemic.

**Design:** Population-based cohort study.

**Setting:** Observational cohort study from 2018 to 2021 using the Information System for Research in Primary Care (SIDIAP) database in Catalonia, Spain.

**Participants:** 4,255,847 individuals aged 18 or older in SIDIAP on 1 March, 2018 with no prior history of anxiety and depressive disorders.

**Primary and secondary outcomes measures:** Incidence of anxiety and depressive disorders prior to COVID-19 (March, 2018 to February, 2020), during the COVID-19 lockdown (March to June, 2020) and post-lockdown periods (from July, 2020 to March, 2021) were calculated. Forecasted rates over COVID-19 periods were estimated using negative binomial regression models based on previous data. The percentage reduction was estimated by comparing forecasted versus observed events, overall and by age, sex and socioeconomic status.

**Results:** The incidence rates per 100,000 person-months of anxiety and depressive disorders were 171.0 (95%CI: 170.2-171.8) and 46.6 (46.2-47.0), respectively, during the pre-lockdown period. We observed an increase of 39.7% (95%PI: 26.5 to 53.3) in incident anxiety diagnoses compared to the expected in March, 2020, followed by a reduction of 16.9% (8.6 to 24.5) during the post-lockdown periods. A reduction of incident depressive disorders occurred during the lockdown and post-lockdown periods (46.6% [38.9 to 53.1] and 23.2% [12.0 to 32.7], respectively). Reductions were higher among adults aged 18 to 34 and individuals living in most deprived areas.

**Conclusions:** The COVID-19 pandemic in Catalonia was associated with an initial increase in anxiety disorders diagnosed in primary care, but a reduction in cases as the pandemic continued. Diagnoses of depressive disorders were lower than expected throughout the pandemic.

**Summary box:** *What is already known on this topic:*

- While previous self-reported studies have provided evidence of increased mental health burden during the initial phase of the COVID-19 pandemic, a number of studies observed that fewer diagnoses were made in primary care settings than would have been expected during the initial stages of the pandemic.
- Population data that examine the impact of COVID-19 on temporal trends of incident cases of common mental health disorders are lacking in Catalonia, Spain.

*What this study adds:*

- This study has quantified the impact of the COVID-19 pandemic on trends of incidence of anxiety and depressive disorders among adults living in Catalonia.
- Reductions in incident cases of anxiety and depressive disorders were higher for young adults and people living in most deprived areas.
- Incident diagnoses of anxiety and depressive disorders have not been fully recovered to what would have been expected.

## INTRODUCTION

The outbreak of the COVID-19 pandemic and the associated control measures have impacted many aspects of people’s lives. In Spain, a national lockdown was implemented on 14 March.^1^ Strict control measures were adopted in order to contain the spread of the virus, limiting mobility with few exceptions such as grocery shopping, health emergencies or essential work. The restrictions were gradually lifted following different de-escalation phases, which started on 28 May, 2020 and ended on 21 June, 2020.^2^ Due to a resurgence of COVID-19 infections, new nationwide measures were implemented on 25 October, 2020, including mobility restrictions and curfew hours, which were extended up to 9 May, 2021.^3^

Studies based on self-reported surveys have provided evidence of elevated rates of anxiety, depression and stress in the initial stages of the pandemic.^4–7^ In Catalonia (a northern region of Spain), self-reported surveys performed in April-May 2020 found increased levels of anxiety and depression during COVID-19 lockdown, especially among women and young adults.^8,9^ Conversely, health care contacts related to mental conditions were substantially reduced in primary care, emergency departments and hospital settings after the lockdown announcement in March, 2020.^10^

Serious concerns have been raised about the long-term and far-reaching mental health impact of the pandemic.^11^ A discrepancy between increasing levels of mental health disorders and reductions in primary care diagnoses during the COVID-19 pandemic may well result in a substantial burden of ill-health, as long delays in diagnosis are associated with negative health outcomes among adults with mood and/or anxiety disorders.^12,13^

To date, most evidence regarding the mental health impact of COVID-19 is from self-report surveys performed during the initial stages of the pandemic. Yet, we do not know whether survey results are mirrored in the rates of recorded incident diagnoses of common mental disorders in primary care, and how they have evolved during the pandemic in Catalonia.

The use of longitudinal studies to monitor rates of mental disorders and to identify gaps in mental health care have been described as an urgent research priority in the response to COVID-19.^14^ In this study, we aim to investigate how incidence trends of anxiety and depressive disorders have been affected by the various stages of the COVID-19 pandemic through the analysis of a large primary care longitudinal dataset representative of the population living in Catalonia, Spain.

## METHODS

### Study design

We conducted a cohort study based on primary care records from 1 March 2018 to 31 March 2021 in Catalonia, Spain. Time trends were assessed in three different periods: 1) the pre-lockdown-19 period from 1 March 2018 to 29 February 2020; 2) the lockdown period from 1 March 2020 to 30 June 2020, and 3) the post-lockdown period, from 1 July 2020 to 31 March 2021. The lockdown period coincides with the home confinement and different phases of de-escalation in Spain. The post-lockdown period was divided into three trimesters, which were broadly aligned with the different stages of the pandemic in Catalonia: 1) easing of restrictions from 1 July, 2020 to 30 September, 2020; 2) implementation of new control measures from 1 October, 2020 to 31 December, 2020; 3) extension of control measures and start of the vaccination campaign from 1 January, 2021 to 31 March, 2021.

### Setting

The Catalan Health Care System dispenses services for 7.5 inhabitants, providing universal health coverage to residents of Catalonia through a tax-based system.^15^ Primary care services in Spain are the first point of contact with healthcare services for the population,^16^ and they carry the major burden of detection, management and treatment of mild mental health problems.^17^

### Study participants and data source

Individual-level data were extracted from the Information System for Research in Primary Care (SIDIAP; www.sidiap.org) database, which captures electronic health records from approximately 80% of the population living in Catalonia and has been shown to be representative of the population in Catalonia in geography, age, and sex.^18^ The database has been mapped to the Observational Medical Outcomes Partnership (OMOP) Common Data Model (CDM).^19^

All individuals aged 18 or older registered in the SIDIAP on 1 March, 2018 were identified. Individuals with less than one year of prior history available were excluded so that study participants had sufficient prior observation time to truly detect incident cases of anxiety and depressive disorders. Individuals with episodes of anxiety or depressive disorders prior to index date (i.e., date of start of the cohort) were excluded. Individuals were observed until the event of interest (anxiety or depressive disorders), until they were transferred or died, or until the study period ended.

### Variables

Individuals’ age and sex were extracted. Information on socioeconomic status (SES) was available through the Mortalidad en áreas pequeñas Españolas y Desigualdades Socioeconómicas y Ambientales (MEDEA) deprivation index, linked to each residential census area of the population.^20^ The deprivation index was only available for urban areas, defined as municipalities with more than 10,000 inhabitants and a population density greater than 150 habitants/km^2^; remaining areas were considered rural areas. The deprivation index is categorized in quintiles, in which the first and fifth quintiles are the least and most deprived, respectively.

The outcomes of the study were incidence of anxiety or depressive disorders. Conditions were identified on the basis of the International Statistical Classification of Diseases and Related Health Problems, Tenth Revision, Clinical Modification (ICD-10-CM) codes of interest. The ICD-10-CM codes included for depressive disorders were F32 (depressive episode) and F33 (recurrent depressive disorder). All descendants were included except the ones referring to episodes in partial or complete remission (F32.4, F32.5 and F33.4). For anxiety disorder the F41 (other anxiety disorder) code and all its descendants were included.

### Statistical methods

We first summarised the socio-demographics characteristics of individuals included in the study, with counts and percentages for categorical variables and median and interquartile ranges (IQR) for continuous variables. We structured data in a monthly time-series format with incident and person-months at risk aggregated with stratification by sex (males and females), age group (18 to 34, 35 to 64, >65 years) and SES (quintiles of MEDEA deprivation index). Incidence rates with 95% CI were calculated for each month and study period by dividing the number of first recorded cases of anxiety and depression by 100,000 person-months at risk, overall and stratified by sex, age group and SES. Incidence Rate Ratios (IRR) with 95% CI were calculated to compare the differences in incidence of each strata of the population during the lockdown period and post-lockdown period (divided into trimesters) compared to the pre-lockdown period.

We used negative binomial regression models to estimate expected monthly incident cases from March, 2020 to March, 2021, using data collected in the pre-lockdown period. To account for possible seasonality and linear trends, we fitted calendar month as a categorical variable and time as a continuous variable. The number of months since the start of the study was considered as the unit of measurement for time.

The estimated number of underdiagnoses were calculated by subtracting the number of expected from the observed diagnoses. Reduction in diagnoses were calculated dividing the number of underdiagnoses by the expected diagnoses, and were estimated monthly and by study period (lockdown and post-lockdown, overall and divided by trimesters). Reduction in diagnoses are expressed as percentages with 95% prediction intervals (PI),^21^ as previously reported in a similar study.^22^

For each month during the COVID-19 study period, observed and expected incident counts were converted to rates using the observed person-month denominator. We plotted monthly expected rates and corresponding 95%PI against the observed rates.

To validate our modelling approach, we developed a negative binomial model based on two years of prior history (from March, 2017 to February, 2019). We forecasted our expected values from March, 2019 to February, 2020, and checked if values predicted from the model fell within the calculated 95%PI. We validated our approach for overall diagnoses and stratifying by sex, age group and SES.

All analyses were performed in R version 4.0.4.

### Patient and public involvement

This research was done without patient involvement. Patients were not invited to comment on the study design and were not consulted to develop patient relevant outcomes or interpret the results. Patients were not invited to contribute to the writing or editing of this document for readability or accuracy.

## RESULTS

Our dataset included 4,255,847 individuals, of whom 2,105,171 (49.5%) were female. The median age of individuals on the index date was 47 years (IQR 35 to 62). Detailed demographic data of the study population is included in the Supplementary Table S1.

During the pre-lockdown period, incidence rate per 100,000 person-months of anxiety or depressive disorders were 171.0 (95%CI 170.2 to 171.8) and 46.6 (46.2 to 47.0), respectively. During the lockdown period, the incidence rate per 100,000 person-months of anxiety disorders increased up to 201.6 (199.4 to 203.9) whereas rates for depressive disorders sharply decreased down to 29.5 per 100,000 person-months (28.6 to 30.3). Incidence rates of anxiety disorders were substantially reduced from July to September, 2020 (142.0 per 100,000 person-months [139.9 to 144.2]), but subsequently increased and exceeded the figures observed before the pandemic during the first trimester of 2021 (149.4 per 100,000 person-months [147.2 to 151.7] from October to December, 2020; 176.1 per 100,000 person-months [173.6 to 178.6] from January to March, 2021). Quarterly incidence rates of depressive disorders progressively recovered after the lockdown period, and achieved the observed figures prior to the pandemic during the first trimester of 2021 (36.0 per 100,000 person-months [34.9 to 37.1] from July to September, 2020; 40.1 per 100,000 person-months [38.9 to 41.2] from October to December, 2020 and 46.6 per 100,000 person-months [45.3 to 47.9] from January to March, 2021) (Figure 1). Detailed data of incidence rates stratified by age, gender and SES during the lockdown and post-lockdown periods is included in the Supplementary Materials.

**Figure 1.**
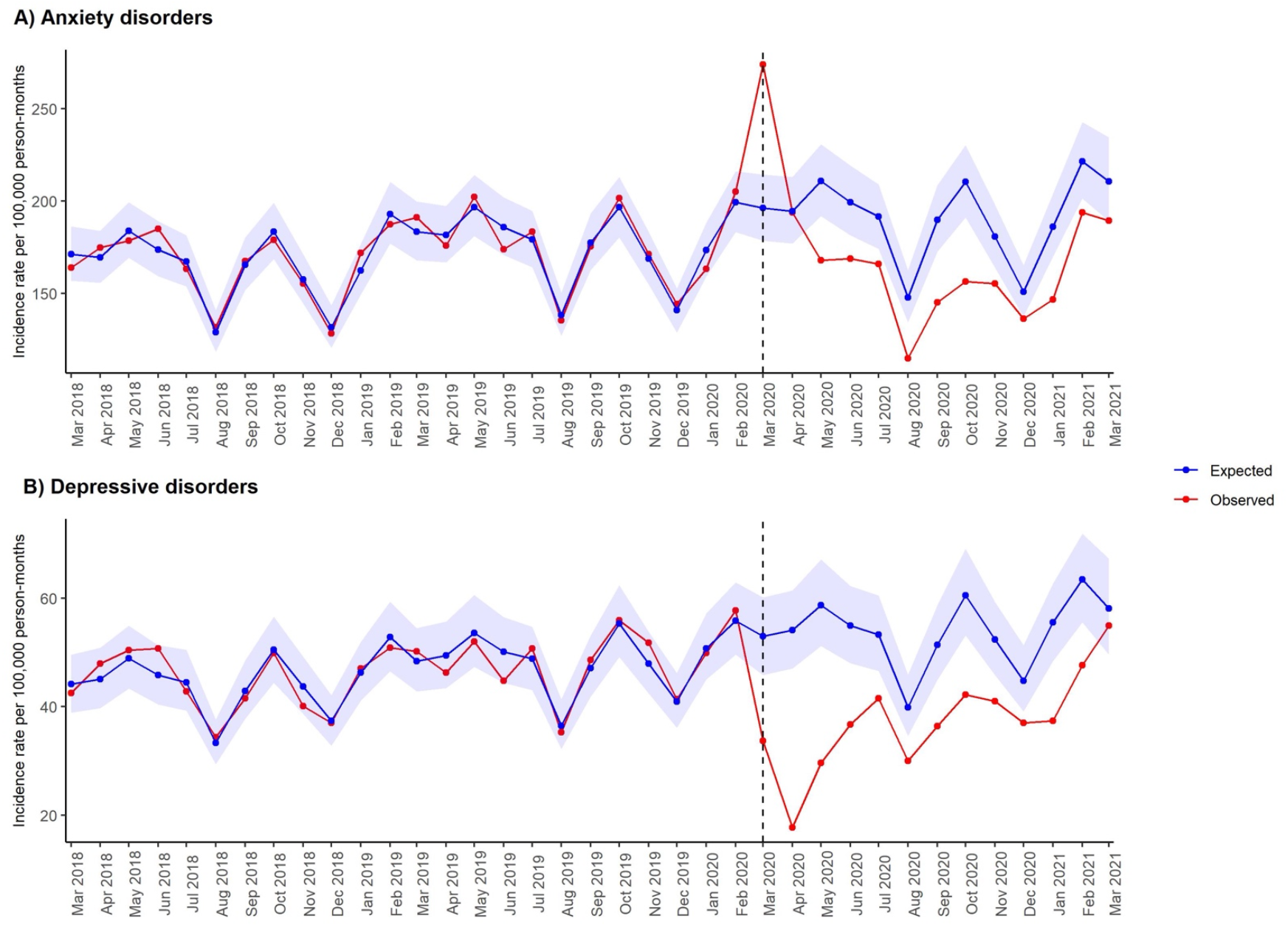
Expected and observed incidence rates of anxiety and depressive disorders in primary care in Catalonia (March, 2018–March, 2021). Points indicate monthly incidence rates of anxiety and depressive disorders. Number of expected cases (95% PI) were estimated with negative binomial models, using data from 1 March, 2018 to 29 February, 2020. Shaded areas in blue represent 95 % PI. Vertical lines show 1 March, 2020.

Incidence Rate Ratios (IRR) comparing the differences in incidence of each strata of the population during the lockdown and post-lockdown periods compared to the pre-lockdown period are shown in Table 1. IRR of anxiety disorders were significantly higher during the lockdown period for all age groups and deprivation quintiles (IRR range 1.06 to 1.37) except for females aged between 18 to 34 years (IRR 1.00 [0.97 to 1.03]). Significant reduction in diagnoses of anxiety disorders were found for all strata during the following two trimesters of 2020. Individuals aged 18 to 34 years or individuals living in rural areas obtained significantly higher IRR during the first trimester of 2021 compared to the pre-lockdown period (ie, IRR 1.13 [1.09 to 1.17] among females aged 18 to 34 years, and 1.07 [1.03-1.11] among people living in rural areas). Significant reductions in diagnoses of depressive disorders were found for all strata during the lockdown and first and second trimester of the post-lockdown period with a few exceptions (ie, IRR range 0.55 to 0.69 during the lockdown period). IRR of depressive disorders increased during the first semester of 2021 (IRR range 0.90 to 1.18), and were significantly higher for females aged 18 to 34 years and males aged 18 to 64 years (IRR 1.15 [1.05 to 1.1.26] among females aged 18 to 34 years, 1.18 [1.06 to 1.32] and 1.06 [1.00 to 1.13] among males aged 18 to 34 and 35 to 64, respectively).

**Table 1.**
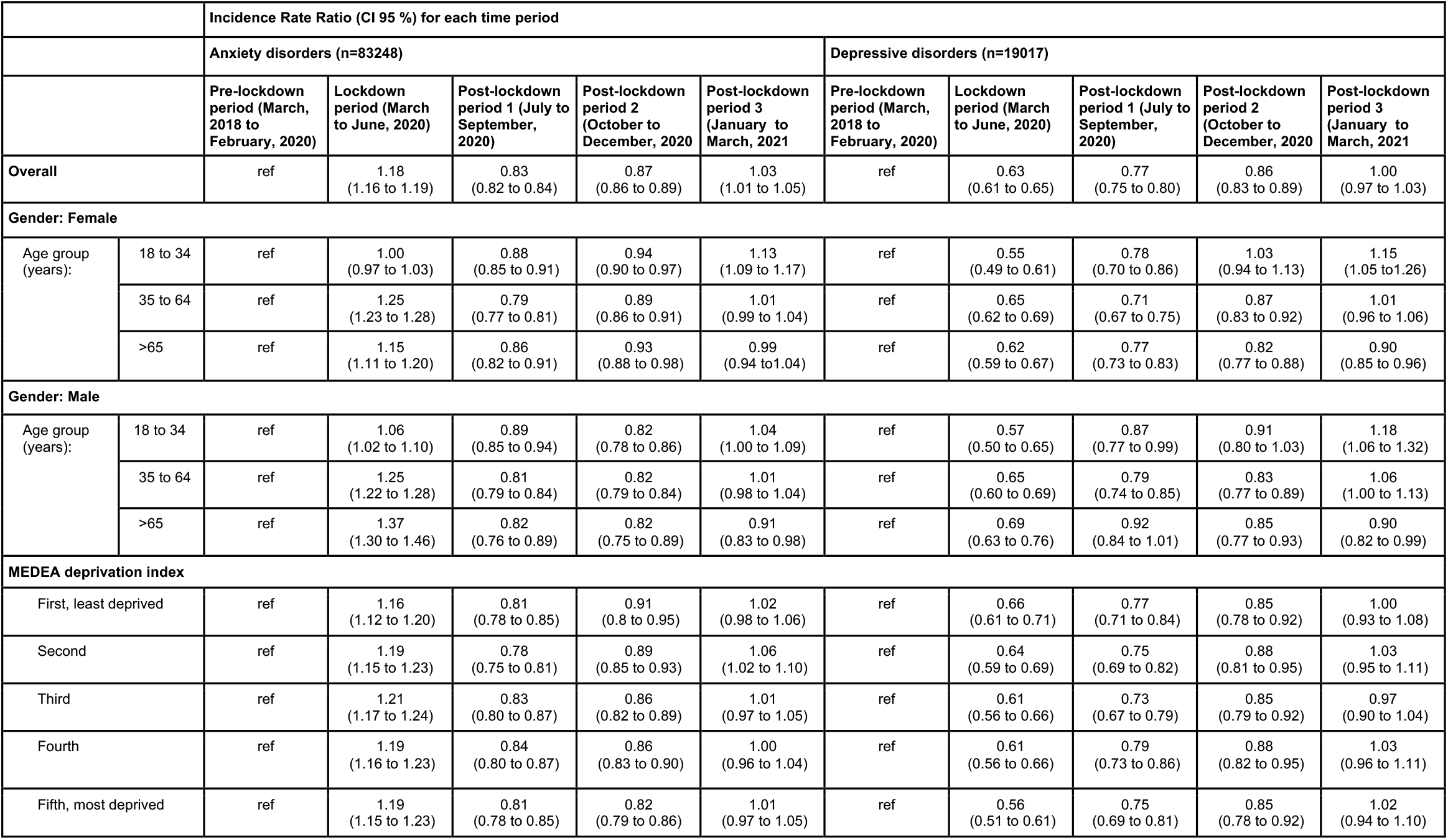
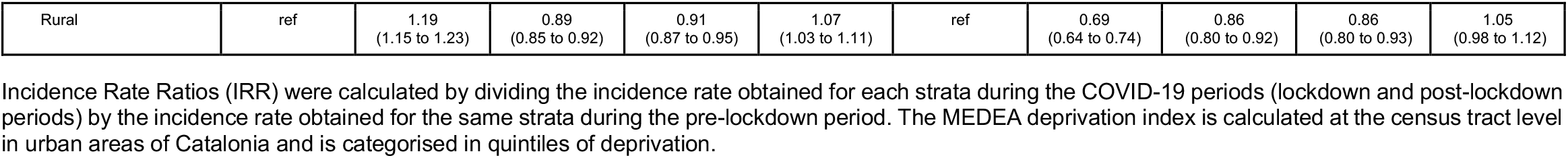
Incidence rate ratio of first recorded cases of anxiety and depressive disorders stratified by gender, age group, gender and MEDEA deprivation index according to the study period.

|  |  | Incidence Rate Ratio (CI 95 %) for each time period |  |  |  |  |  |  |  |  |  |
| --- | --- | --- | --- | --- | --- | --- | --- | --- | --- | --- | --- |
|  |  | Anxiety disorders (n=83248) |  |  |  |  | Depressive disorders (n=19017) |  |  |  |  |
|  |  | Pre-lockdown period (March, 2018 to February, 2020) | Lockdown period (March to June, 2020) | Post-lockdown period 1 (July to September, 2020) | Post-lockdown period 2 (October to December, 2020) | Post-lockdown period 3 (January to March, 2021) | Pre-lockdown period (March, 2018 to February, 2020) | Lockdown period (March to June, 2020) | Post-lockdown period 1 (July to September, 2020) | Post-lockdown period 2 (October to December, 2020) | Post-lockdown period 3 (January to March, 2021) |
| Overall |  | ref | 1.18<br>(1.16 to 1.19) | 0.83<br>(0.82 to 0.84) | 0.87<br>(0.86 to 0.89) | 1.03<br>(1.01 to 1.05) | ref | 0.63<br>(0.61 to 0.65) | 0.77<br>(0.75 to 0.80) | 0.86<br>(0.83 to 0.89) | 1.00<br>(0.97 to 1.03) |
| Gender: Female |  |  |  |  |  |  |  |  |  |  |  |
| Age group (years): | 18 to 34 | ref | 1.00<br>(0.97 to 1.03) | 0.88<br>(0.85 to 0.91) | 0.94<br>(0.90 to 0.97) | 1.13<br>(1.09 to 1.17) | ref | 0.55<br>(0.49 to 0.61) | 0.78<br>(0.70 to 0.86) | 1.03<br>(0.94 to 1.13) | 1.15<br>(1.05 to 1.26) |
|  | 35 to 64 | ref | 1.25<br>(1.23 to 1.28) | 0.79<br>(0.77 to 0.81) | 0.89<br>(0.86 to 0.91) | 1.01<br>(0.99 to 1.04) | ref | 0.65<br>(0.62 to 0.69) | 0.71<br>(0.67 to 0.75) | 0.87<br>(0.83 to 0.92) | 1.01<br>(0.96 to 1.06) |
|  | >65 | ref | 1.15<br>(1.11 to 1.20) | 0.86<br>(0.82 to 0.91) | 0.93<br>(0.88 to 0.98) | 0.99<br>(0.94 to 1.04) | ref | 0.62<br>(0.59 to 0.67) | 0.77<br>(0.73 to 0.83) | 0.82<br>(0.77 to 0.88) | 0.90<br>(0.85 to 0.96) |
| Gender: Male |  |  |  |  |  |  |  |  |  |  |  |
| Age group (years): | 18 to 34 | ref | 1.06<br>(1.02 to 1.10) | 0.89<br>(0.85 to 0.94) | 0.82<br>(0.78 to 0.86) | 1.04<br>(1.00 to 1.09) | ref | 0.57<br>(0.50 to 0.65) | 0.87<br>(0.77 to 0.99) | 0.91<br>(0.80 to 1.03) | 1.18<br>(1.06 to 1.32) |
|  | 35 to 64 | ref | 1.25<br>(1.22 to 1.28) | 0.81<br>(0.79 to 0.84) | 0.82<br>(0.79 to 0.84) | 1.01<br>(0.98 to 1.04) | ref | 0.65<br>(0.60 to 0.69) | 0.79<br>(0.74 to 0.85) | 0.83<br>(0.77 to 0.89) | 1.06<br>(1.00 to 1.13) |
|  | >65 | ref | 1.37<br>(1.30 to 1.46) | 0.82<br>(0.76 to 0.89) | 0.82<br>(0.75 to 0.89) | 0.91<br>(0.83 to 0.98) | ref | 0.69<br>(0.63 to 0.76) | 0.92<br>(0.84 to 1.01) | 0.85<br>(0.77 to 0.93) | 0.90<br>(0.82 to 0.99) |
| MEDEA deprivation index |  |  |  |  |  |  |  |  |  |  |  |
| First, least deprived |  | ref | 1.16<br>(1.12 to 1.20) | 0.81<br>(0.78 to 0.85) | 0.91<br>(0.8 to 0.95) | 1.02<br>(0.98 to 1.06) | ref | 0.66<br>(0.61 to 0.71) | 0.77<br>(0.71 to 0.84) | 0.85<br>(0.78 to 0.92) | 1.00<br>(0.93 to 1.08) |
| Second |  | ref | 1.19<br>(1.15 to 1.23) | 0.78<br>(0.75 to 0.81) | 0.89<br>(0.85 to 0.93) | 1.06<br>(1.02 to 1.10) | ref | 0.64<br>(0.59 to 0.69) | 0.75<br>(0.69 to 0.82) | 0.88<br>(0.81 to 0.95) | 1.03<br>(0.95 to 1.11) |
| Third |  | ref | 1.21<br>(1.17 to 1.24) | 0.83<br>(0.80 to 0.87) | 0.86<br>(0.82 to 0.89) | 1.01<br>(0.97 to 1.05) | ref | 0.61<br>(0.56 to 0.66) | 0.73<br>(0.67 to 0.79) | 0.85<br>(0.79 to 0.92) | 0.97<br>(0.90 to 1.04) |
| Fourth |  | ref | 1.19<br>(1.16 to 1.23) | 0.84<br>(0.80 to 0.87) | 0.86<br>(0.83 to 0.90) | 1.00<br>(0.96 to 1.04) | ref | 0.61<br>(0.56 to 0.66) | 0.79<br>(0.73 to 0.86) | 0.88<br>(0.82 to 0.95) | 1.03<br>(0.96 to 1.11) |
| Fifth, most deprived |  | ref | 1.19<br>(1.15 to 1.23) | 0.81<br>(0.78 to 0.85) | 0.82<br>(0.79 to 0.86) | 1.01<br>(0.97 to 1.05) | ref | 0.56<br>(0.51 to 0.61) | 0.75<br>(0.69 to 0.81) | 0.85<br>(0.78 to 0.92) | 1.02<br>(0.94 to 1.10) |

|  |  |  |  |  |  |  |  |  |  |  |
| --- | --- | --- | --- | --- | --- | --- | --- | --- | --- | --- |
| Rural | ref | 1.19<br>(1.15 to 1.23) | 0.89<br>(0.85 to 0.92) | 0.91<br>(0.87 to 0.95) | 1.07<br>(1.03 to 1.11) | ref | 0.69<br>(0.64 to 0.74) | 0.86<br>(0.80 to 0.92) | 0.86<br>(0.80 to 0.93) | 1.05<br>(0.98 to 1.12) |

Incidence Rate Ratios (IRR) were calculated by dividing the incidence rate obtained for each strata during the COVID-19 periods (lockdown and post-lockdown periods) by the incidence rate obtained for the same strata during the pre-lockdown period. The MEDEA deprivation index is calculated at the census tract level in urban areas of Catalonia and is categorised in quintiles of deprivation.

Incident diagnoses of anxiety disorders showed a sudden peak at the onset of the lockdown measures in Catalonia, and accounted for an increase of 39.7% (95%PI 26.5 to 53.3) new diagnoses compared to the expected in March, 2020 (Figure 1A). This peak was particularly pronounced among individuals aged 18 to 64 years and individuals living in most deprived areas (Figure 2A and Figure 3A). Compared to the younger groups, males aged 65 or older showed a lower increase of incident cases of anxiety disorders from April to May, 2020, which was much more subtle among their female peer group (Figure 2A).

**Figure 2.**
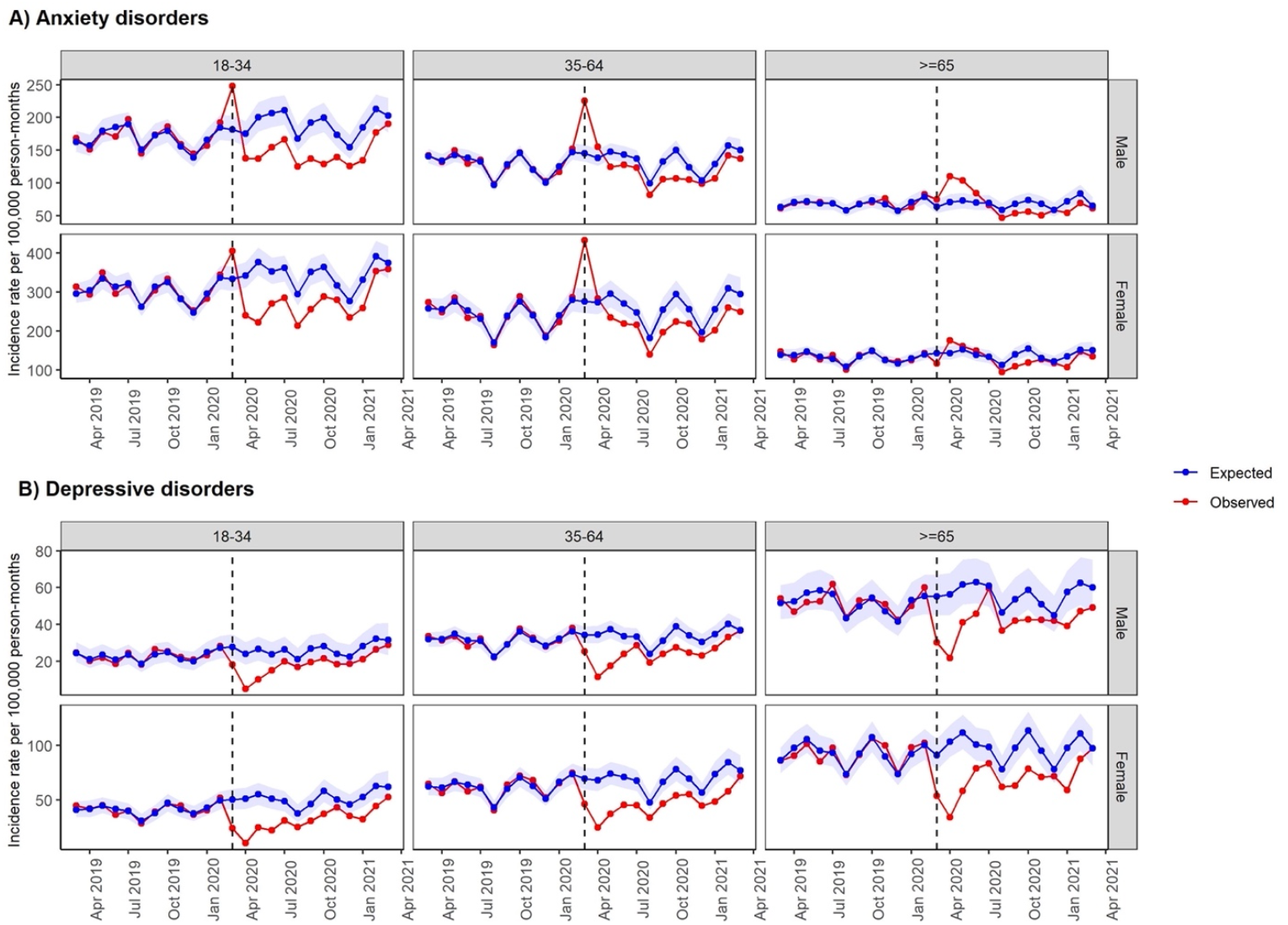
Expected and observed incidence rates of anxiety and depressive disorders stratified by age group and gender in primary care in Catalonia (March, 2019–March, 2021). Points indicate monthly incidence rates of anxiety and depressive disorders. Number of expected cases (95% PI) were estimated with negative binomial models, using data from 1 March, 2018 to 29 February, 2020. Shaded areas in blue represent 95 % PI. Vertical lines show 1 March, 2020.

**Figure 3.**
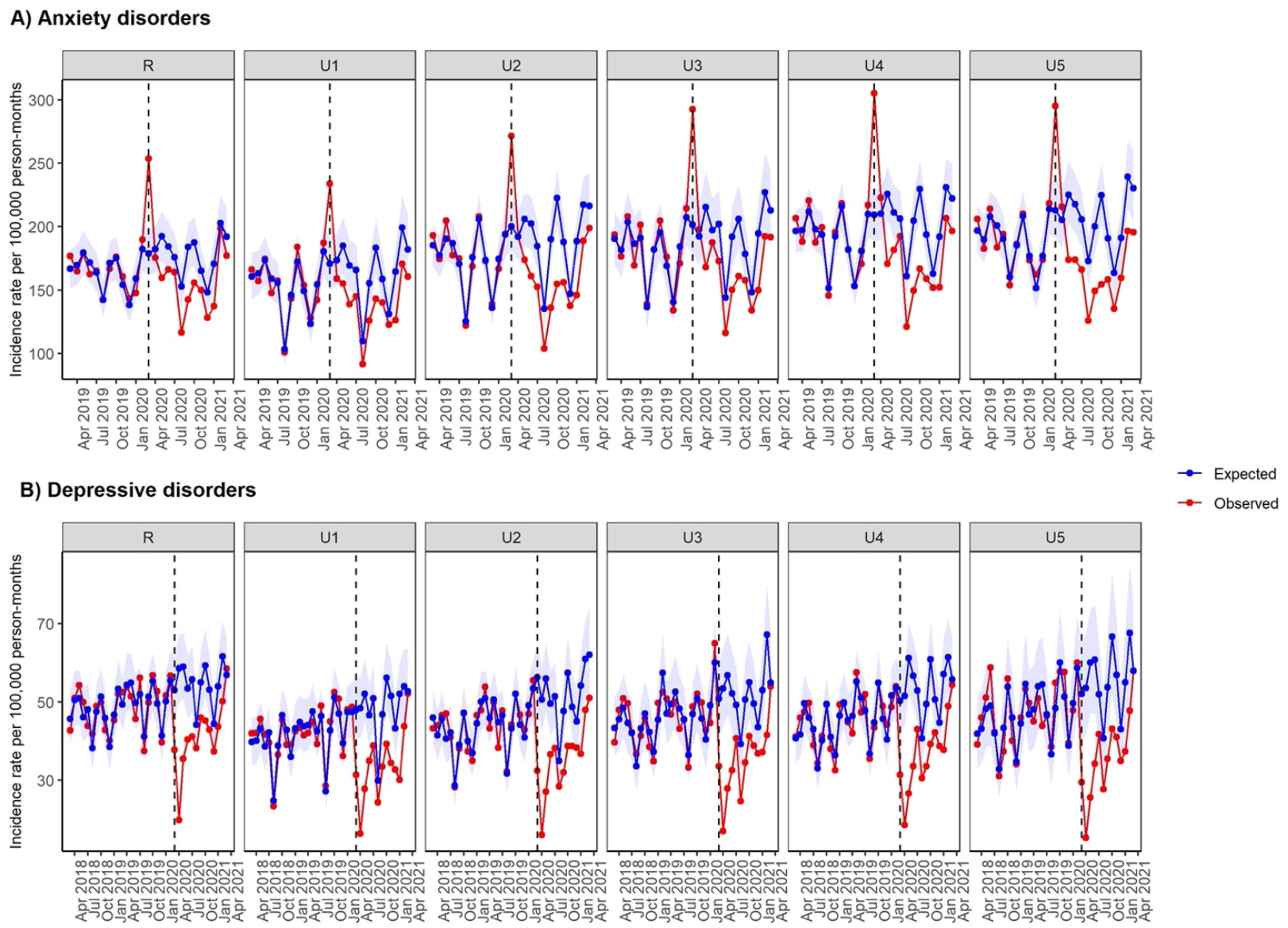
Expected and observed incidence rates of anxiety and depressive disorders stratified by MEDEA deprivation index in primary care in Catalonia (March, 2019–March, 2021). Points indicate monthly incidence rates of anxiety and depressive disorders. Number of expected cases (95% PI) were estimated with negative binomial models, using data from 1 March, 2018 to 29 February, 2020. Shaded areas in blue represent 95 % PI. Vertical lines show 1 March, 2020. The MEDEA deprivation index is calculated at the census tract level in urban areas of Catalonia and is categorised in quintiles of deprivation. It also includes a rural category for individuals living in rural areas.

The estimated number of underdiagnoses and the percentage of reduction are presented stratified by age, sex and SES in Table 2. An increase of diagnoses of anxiety disorders was found during the lockdown period for all stratas except from individuals aged 18 to 34 years, as new diagnoses of anxiety disorders among this subgroup were lower than expected from April, 2020 to February, 2021 (Figure 2A; Table 2). The increase of diagnoses was followed by a decrease of 19.4% (95%PI 11.4 to 26.6) compared to the expected from July to August, 2020. This percentage was progressively reduced during the following two trimesters (17.3% [9.3 to 24.8] from October to December, 2020; 14.3 % [5.6 to 22.6] from January to March, 2021). The reduction of incident cases of post-lockdown period was 16.9% (8.6 to 24.5) (Table 2).

**Table 2.**
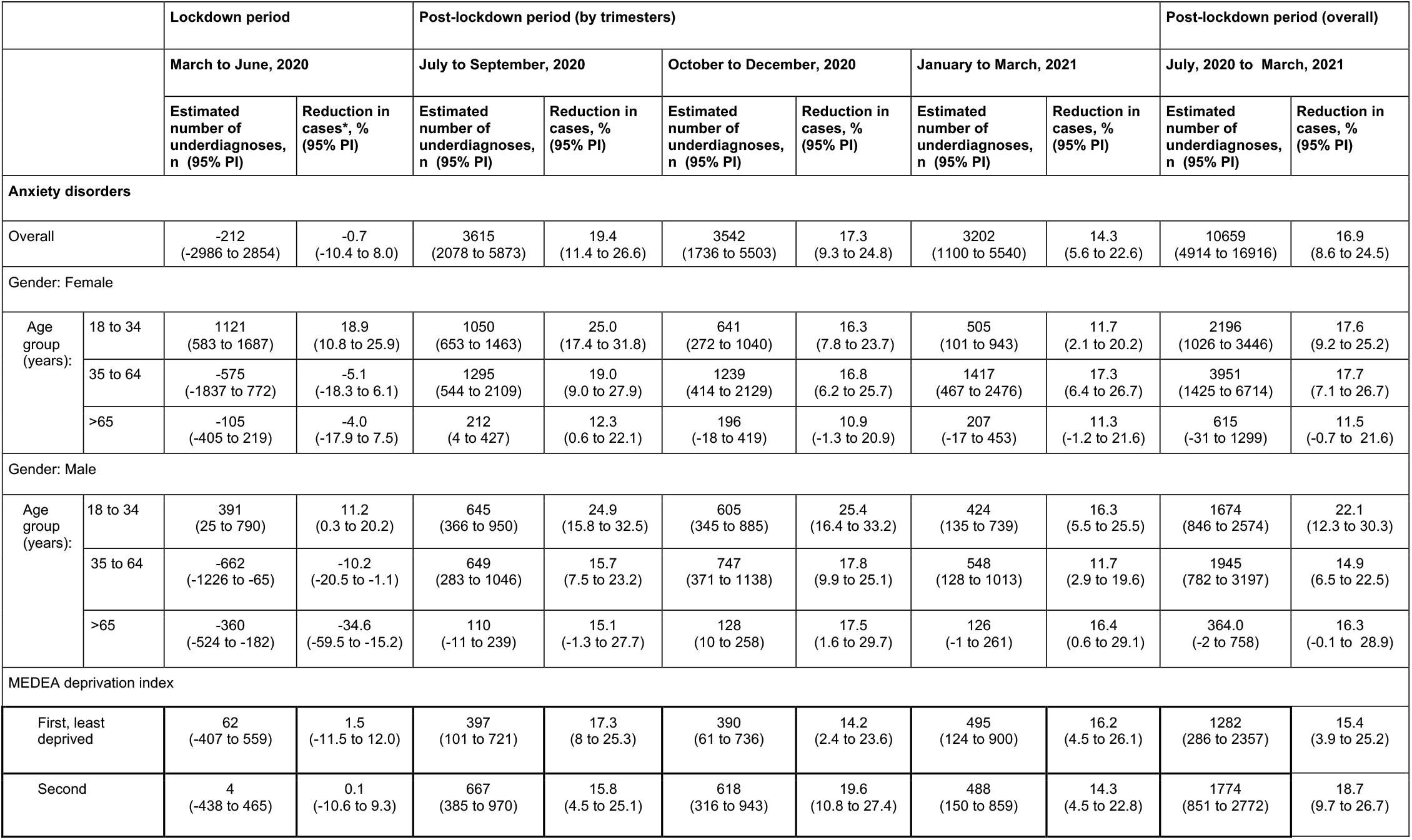
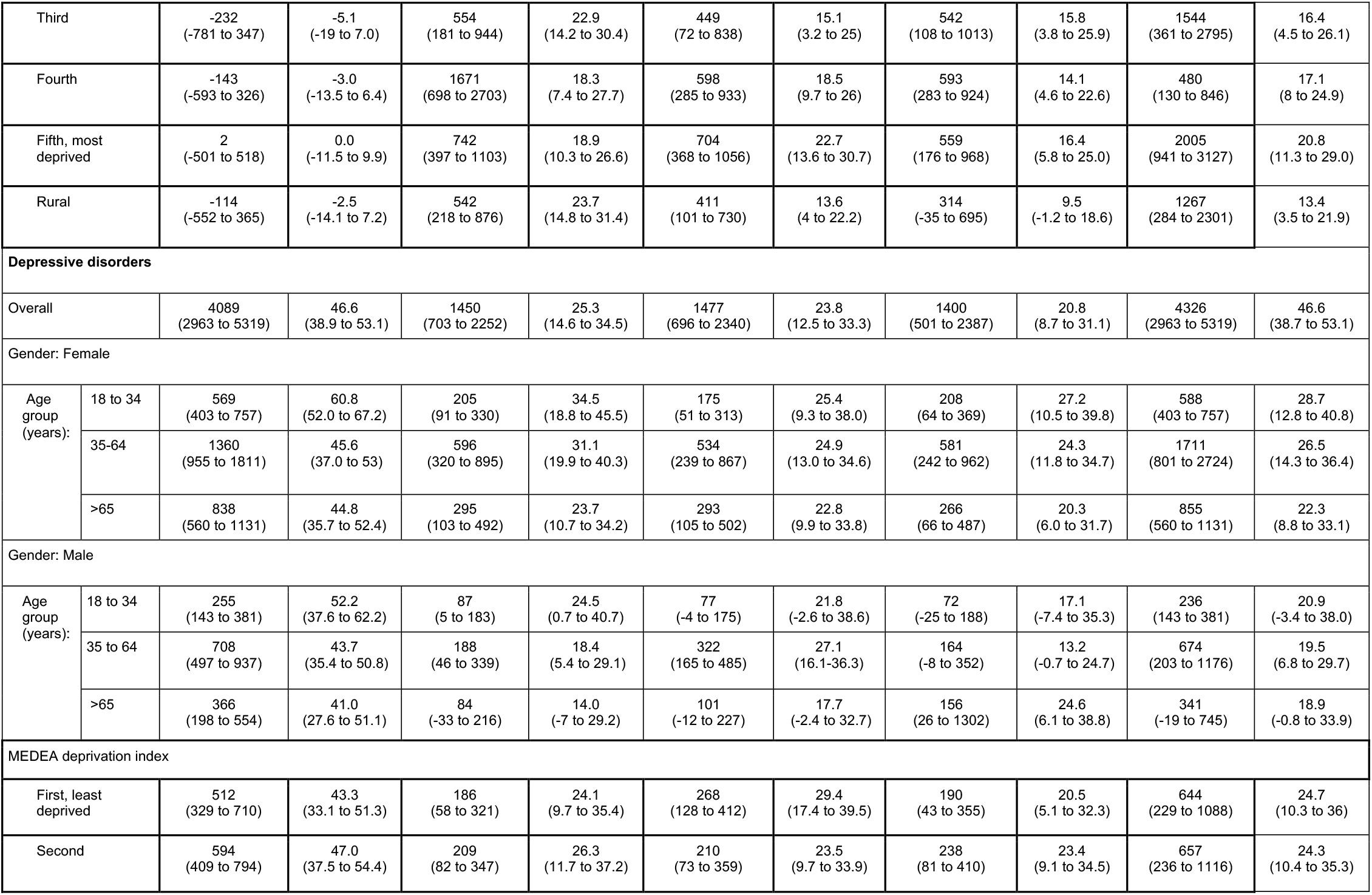
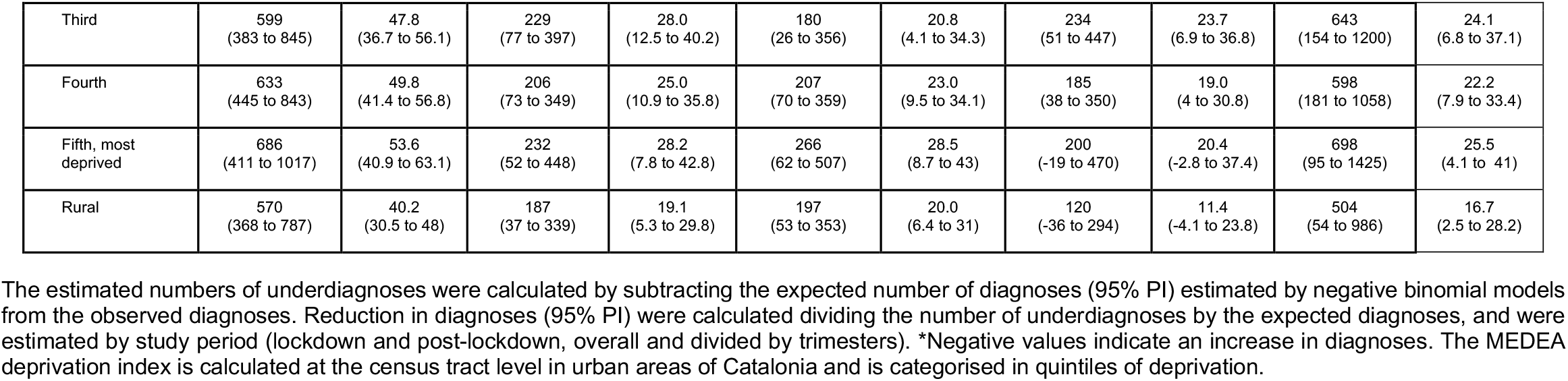
Estimated number of underdiagnosed of anxiety and depressive disorders and percentage (%) of reduction compared with expected, by lockdown and post-lockdown period.

|  |  | Lockdown period |  | Post-lockdown period (by trimesters) |  |  |  |  |  | Post-lockdown period (overall) |  |
| --- | --- | --- | --- | --- | --- | --- | --- | --- | --- | --- | --- |
|  |  | March to June, 2020 |  | July to September, 2020 |  | October to December, 2020 |  | January to March, 2021 |  | July, 2020 to March, 2021 |  |
|  |  | Estimated number of underdiagnoses, n (95% PI) | Reduction in cases*, % (95% PI) | Estimated number of underdiagnoses, n (95% PI) | Reduction in cases, % (95% PI) | Estimated number of underdiagnoses, n (95% PI) | Reduction in cases, % (95% PI) | Estimated number of underdiagnoses, n (95% PI) | Reduction in cases, % (95% PI) | Estimated number of underdiagnoses, n (95% PI) | Reduction in cases, % (95% PI) |
| **Anxiety disorders** |  |  |  |  |  |  |  |  |  |  |  |
| Overall |  | -212<br>(-2986 to 2854) | -0.7<br>(-10.4 to 8.0) | 3615<br>(2078 to 5873) | 19.4<br>(11.4 to 26.6) | 3542<br>(1736 to 5503) | 17.3<br>(9.3 to 24.8) | 3202<br>(1100 to 5540) | 14.3<br>(5.6 to 22.6) | 10659<br>(4914 to 16916) | 16.9<br>(8.6 to 24.5) |
| Gender: Female |  |  |  |  |  |  |  |  |  |  |  |
| Age group (years): | 18 to 34 | 1121<br>(583 to 1687) | 18.9<br>(10.8 to 25.9) | 1050<br>(653 to 1463) | 25.0<br>(17.4 to 31.8) | 641<br>(272 to 1040) | 16.3<br>(7.8 to 23.7) | 505<br>(101 to 943) | 11.7<br>(2.1 to 20.2) | 2196<br>(1026 to 3446) | 17.6<br>(9.2 to 25.2) |
|  | 35 to 64 | -575<br>(-1837 to 772) | -5.1<br>(-18.3 to 6.1) | 1295<br>(544 to 2109) | 19.0<br>(9.0 to 27.9) | 1239<br>(414 to 2129) | 16.8<br>(6.2 to 25.7) | 1417<br>(467 to 2476) | 17.3<br>(6.4 to 26.7) | 3951<br>(1425 to 6714) | 17.7<br>(7.1 to 26.7) |
|  | >65 | -105<br>(-405 to 219) | -4.0<br>(-17.9 to 7.5) | 212<br>(4 to 427) | 12.3<br>(0.6 to 22.1) | 196<br>(-18 to 419) | 10.9<br>(-1.3 to 20.9) | 207<br>(-17 to 453) | 11.3<br>(-1.2 to 21.6) | 615<br>(-31 to 1299) | 11.5<br>(-0.7 to 21.6) |
| Gender: Male |  |  |  |  |  |  |  |  |  |  |  |
| Age group (years): | 18 to 34 | 391<br>(25 to 790) | 11.2<br>(0.3 to 20.2) | 645<br>(366 to 950) | 24.9<br>(15.8 to 32.5) | 605<br>(345 to 885) | 25.4<br>(16.4 to 33.2) | 424<br>(135 to 739) | 16.3<br>(5.5 to 25.5) | 1674<br>(846 to 2574) | 22.1<br>(12.3 to 30.3) |
|  | 35 to 64 | -662<br>(-1226 to -65) | -10.2<br>(-20.5 to -1.1) | 649<br>(283 to 1046) | 15.7<br>(7.5 to 23.2) | 747<br>(371 to 1138) | 17.8<br>(9.9 to 25.1) | 548<br>(128 to 1013) | 11.7<br>(2.9 to 19.6) | 1945<br>(782 to 3197) | 14.9<br>(6.5 to 22.5) |
|  | >65 | -360<br>(-524 to -182) | -34.6<br>(-59.5 to -15.2) | 110<br>(-11 to 239) | 15.1<br>(-1.3 to 27.7) | 128<br>(10 to 258) | 17.5<br>(1.6 to 29.7) | 126<br>(-1 to 261) | 16.4<br>(0.6 to 29.1) | 364.0<br>(-2 to 758) | 16.3<br>(-0.1 to 28.9) |
| MEDEA deprivation index |  |  |  |  |  |  |  |  |  |  |  |
| First, least deprived |  | 62<br>(-407 to 559) | 1.5<br>(-11.5 to 12.0) | 397<br>(101 to 721) | 17.3<br>(8 to 25.3) | 390<br>(61 to 736) | 14.2<br>(2.4 to 23.6) | 495<br>(124 to 900) | 16.2<br>(4.5 to 26.1) | 1282<br>(286 to 2357) | 15.4<br>(3.9 to 25.2) |
| Second |  | 4<br>(-438 to 465) | 0.1<br>(-10.6 to 9.3) | 667<br>(385 to 970) | 15.8<br>(4.5 to 25.1) | 618<br>(316 to 943) | 19.6<br>(10.8 to 27.4) | 488<br>(150 to 859) | 14.3<br>(4.5 to 22.8) | 1774<br>(851 to 2772) | 18.7<br>(9.7 to 26.7) |
| Third |  | -232<br>(-781 to 347) | -5.1<br>(-19 to 7.0) | 554<br>(181 to 944) | 22.9<br>(14.2 to 30.4) | 449<br>(72 to 838) | 15.1<br>(3.2 to 25) | 542<br>(108 to 1013) | 15.8<br>(3.8 to 25.9) | 1544<br>(361 to 2795) | 16.4<br>(4.5 to 26.1) |
| Fourth |  | -143<br>(-593 to 326) | -3.0<br>(-13.5 to 6.4) | 1671<br>(698 to 2703) | 18.3<br>(7.4 to 27.7) | 598<br>(285 to 933) | 18.5<br>(9.7 to 26) | 593<br>(283 to 924) | 14.1<br>(4.6 to 22.6) | 480<br>(130 to 846) | 17.1<br>(8 to 24.9) |
| Fifth, most<br>deprived |  | 2<br>(-501 to 518) | 0.0<br>(-11.5 to 9.9) | 742<br>(397 to 1103) | 18.9<br>(10.3 to 26.6) | 704<br>(368 to 1056) | 22.7<br>(13.6 to 30.7) | 559<br>(176 to 968) | 16.4<br>(5.8 to 25.0) | 2005<br>(941 to 3127) | 20.8<br>(11.3 to 29.0) |
| Rural |  | -114<br>(-552 to 365) | -2.5<br>(-14.1 to 7.2) | 542<br>(218 to 876) | 23.7<br>(14.8 to 31.4) | 411<br>(101 to 730) | 13.6<br>(4 to 22.2) | 314<br>(-35 to 695) | 9.5<br>(-1.2 to 18.6) | 1267<br>(284 to 2301) | 13.4<br>(3.5 to 21.9) |
| **Depressive disorders** |  |  |  |  |  |  |  |  |  |  |  |
| Overall |  | 4089<br>(2963 to 5319) | 46.6<br>(38.9 to 53.1) | 1450<br>(703 to 2252) | 25.3<br>(14.6 to 34.5) | 1477<br>(696 to 2340) | 23.8<br>(12.5 to 33.3) | 1400<br>(501 to 2387) | 20.8<br>(8.7 to 31.1) | 4326<br>(2963 to 5319) | 46.6<br>(38.7 to 53.1) |
| **Gender: Female** |  |  |  |  |  |  |  |  |  |  |  |
| Age<br>group<br>(years): | 18 to 34 | 569<br>(403 to 757) | 60.8<br>(52.0 to 67.2) | 205<br>(91 to 330) | 34.5<br>(18.8 to 45.5) | 175<br>(51 to 313) | 25.4<br>(9.3 to 38.0) | 208<br>(64 to 369) | 27.2<br>(10.5 to 39.8) | 588<br>(403 to 757) | 28.7<br>(12.8 to 40.8) |
|  | 35-64 | 1360<br>(955 to 1811) | 45.6<br>(37.0 to 53) | 596<br>(320 to 895) | 31.1<br>(19.9 to 40.3) | 534<br>(239 to 867) | 24.9<br>(13.0 to 34.6) | 581<br>(242 to 962) | 24.3<br>(11.8 to 34.7) | 1711<br>(801 to 2724) | 26.5<br>(14.3 to 36.4) |
|  | >65 | 838<br>(560 to 1131) | 44.8<br>(35.7 to 52.4) | 295<br>(103 to 492) | 23.7<br>(10.7 to 34.2) | 293<br>(105 to 502) | 22.8<br>(9.9 to 33.8) | 266<br>(66 to 487) | 20.3<br>(6.0 to 31.7) | 855<br>(560 to 1131) | 22.3<br>(8.8 to 33.1) |
| **Gender: Male** |  |  |  |  |  |  |  |  |  |  |  |
| Age<br>group<br>(years): | 18 to 34 | 255<br>(143 to 381) | 52.2<br>(37.6 to 62.2) | 87<br>(5 to 183) | 24.5<br>(0.7 to 40.7) | 77<br>(-4 to 175) | 21.8<br>(-2.6 to 38.6) | 72<br>(-25 to 188) | 17.1<br>(-7.4 to 35.3) | 236<br>(143 to 381) | 20.9<br>(-3.4 to 38.0) |
|  | 35 to 64 | 708<br>(497 to 937) | 43.7<br>(35.4 to 50.8) | 188<br>(46 to 339) | 18.4<br>(5.4 to 29.1) | 322<br>(165 to 485) | 27.1<br>(16.1-36.3) | 164<br>(-8 to 352) | 13.2<br>(-0.7 to 24.7) | 674<br>(203 to 1176) | 19.5<br>(6.8 to 29.7) |
|  | >65 | 366<br>(198 to 554) | 41.0<br>(27.6 to 51.1) | 84<br>(-33 to 216) | 14.0<br>(-7 to 29.2) | 101<br>(-12 to 227) | 17.7<br>(-2.4 to 32.7) | 156<br>(26 to 1302) | 24.6<br>(6.1 to 38.8) | 341<br>(-19 to 745) | 18.9<br>(-0.8 to 33.9) |
| **MEDEA deprivation index** |  |  |  |  |  |  |  |  |  |  |  |
| First, least<br>deprived |  | 512<br>(329 to 710) | 43.3<br>(33.1 to 51.3) | 186<br>(58 to 321) | 24.1<br>(9.7 to 35.4) | 268<br>(128 to 412) | 29.4<br>(17.4 to 39.5) | 190<br>(43 to 355) | 20.5<br>(5.1 to 32.3) | 644<br>(229 to 1088) | 24.7<br>(10.3 to 36) |
| Second |  | 594<br>(409 to 794) | 47.0<br>(37.5 to 54.4) | 209<br>(82 to 347) | 26.3<br>(11.7 to 37.2) | 210<br>(73 to 359) | 23.5<br>(9.7 to 33.9) | 238<br>(81 to 410) | 23.4<br>(9.1 to 34.5) | 657<br>(236 to 1116) | 24.3<br>(10.4 to 35.3) |

|  |  |  |  |  |  |  |  |  |  |  |
| --- | --- | --- | --- | --- | --- | --- | --- | --- | --- | --- |
| Third | 599<br>(383 to 845) | 47.8<br>(36.7 to 56.1) | 229<br>(77 to 397) | 28.0<br>(12.5 to 40.2) | 180<br>(26 to 356) | 20.8<br>(4.1 to 34.3) | 234<br>(51 to 447) | 23.7<br>(6.9 to 36.8) | 643<br>(154 to 1200) | 24.1<br>(6.8 to 37.1) |
| Fourth | 633<br>(445 to 843) | 49.8<br>(41.4 to 56.8) | 206<br>(73 to 349) | 25.0<br>(10.9 to 35.8) | 207<br>(70 to 359) | 23.0<br>(9.5 to 34.1) | 185<br>(38 to 350) | 19.0<br>(4 to 30.8) | 598<br>(181 to 1058) | 22.2<br>(7.9 to 33.4) |
| Fifth, most<br>deprived | 686<br>(411 to 1017) | 53.6<br>(40.9 to 63.1) | 232<br>(52 to 448) | 28.2<br>(7.8 to 42.8) | 266<br>(62 to 507) | 28.5<br>(8.7 to 43) | 200<br>(-19 to 470) | 20.4<br>(-2.8 to 37.4) | 698<br>(95 to 1425) | 25.5<br>(4.1 to 41) |
| Rural | 570<br>(368 to 787) | 40.2<br>(30.5 to 48) | 187<br>(37 to 339) | 19.1<br>(5.3 to 29.8) | 197<br>(53 to 353) | 20.0<br>(6.4 to 31) | 120<br>(-36 to 294) | 11.4<br>(-4.1 to 23.8) | 504<br>(54 to 986) | 16.7<br>(2.5 to 28.2) |

The estimated numbers of underdiagnoses were calculated by subtracting the expected number of diagnoses (95% PI) estimated by negative binomial models from the observed diagnoses. Reduction in diagnoses (95% PI) were calculated dividing the number of underdiagnoses by the expected diagnoses, and were estimated by study period (lockdown and post-lockdown, overall and divided by trimesters). \*Negative values indicate an increase in diagnoses. The MEDEA deprivation index is calculated at the census tract level in urban areas of Catalonia and is categorised in quintiles of deprivation.

Reductions in incident cases of anxiety disorders were greater in females during the lockdown period and were slightly higher in males during the post-lockdown period (lockdown: 2.2% [−9.3 to 12.0] in females vs −5.7% [−17.4 to 4.5] in males; post-lockdown: 16.8% [6.7 to 25.5] in females vs 17.5% [8.0 to 25.8] in males; data not shown). Individuals aged 18 to 34 years had greater reductions of incident diagnoses of anxiety disorders during lockdown and post-lockdown periods. All age groups showed a decrease in the percentage of reduction comparing the first trimester of 2021 with the trimester following the lockdown end, except from males aged 65 or over (Table 2).

New cases of depressive disorders were substantially reduced for all age groups compared to the expected at the onset of the lockdown measures, accounting for a reduction of 67.2% (95%PI 62.5 to 71.3) compared to the expected in April, 2020 (Figure 2B). Overall monthly incidence rates remained under the 95% PI from March, 2020 to February, 2021 (Figure 1B). We estimated that the difference between expected and observed diagnoses accounted for a reduction of 46.6% (38.7 to 53.1) during the lockdown period compared to the expected. This reduction was substantially decreased to 25.3% (14.6 to 34.5) from July, 2020 to September, 2020, and was progressively diminished during the following two trimesters (23.8% [12.5 to 33.3] from October, 2020 to December, 2020; 20.8% [8.7 to 31.1] from January, 2021 to March, 2021). Considering the three trimesters combined, the overall reduction of incident cases of post-lockdown period was 23.2% (12.0 to 32.7) (Table 2).

Reductions in diagnoses of depressive disorders were greater in females than in males during the lockdown and post-lockdown periods (lockdown: 47.8% [38.8 to 55.0] in females vs 44.3% [33.4 to 52.8] in males; post-lockdown: 25.6% [12.0 to 36.1] in females vs 19.6% [3.1 to 32.6] in males, data not shown). All age groups showed a marked decline in the percentage of reduction of incident cases of depressive disorders during the trimester following the lockdown end (Table 2). Reductions were progressively diminished over the following trimester for all age groups with a few exceptions. Males aged 35 to 64 years and 65 years or above experienced a subsequent increase in the reduction of diagnosis of depressive disorders. This increase was limited to the last trimester of 2020 among males aged 35 to 64 years, whereas progressively increased among males aged 65 or above, reaching a 10.6 % absolute increase comparing the first quarter of 2021 with the trimester following the lockdown end. Reductions among females were progressively diminished over time, but slightly increased among females aged 18 to 34 years from January to March 2021. While reductions among individuals aged 18 to 34 and 35 to 64 years were similar from July, 2020 to March, 2021, individuals aged 18 to 34 years were the age group with higher reductions during the lockdown and post-lockdown periods (Table 2).

Most deprived urban areas accounted for higher reductions of incident diagnoses of anxiety disorders (during the lockdown period) and depressive disorders (during the lockdown and post-lockdown periods) (Table 2). Reductions were lower in rural areas compared to urban areas for both periods, but were particularly high for anxiety disorders from July to September, 2020 (23.7 % [95%PI 14.8 to 31.4]). Disparities in the reductions of incident diagnoses of anxiety and depressive disorders between the least and most deprived urban areas were reduced during the first quarter of 2021, and obtained similar figures from January to March, 2021 (ie, reduction of cases of anxiety disorders of 16.2% in least deprived areas vs 16.4% in most deprived areas). Reductions of diagnoses of anxiety and depressive disorders decreased during the first quarter of 2021 compared to the first trimester after the lockdown for urban and rural areas (Figure 3).

Our validation approach obtainined consistent performances overall and stratifying by sex, age group and SES. Figures of monthly observed expected incidence rates and corresponding 95%PI from March, 2017 to February, 2020 are included in the Supplementary Material.

## DISCUSSION

In this large population-based cohort study, we observed large variations in the incidence of anxiety and depression disorders diagnosed in primary care in Catalonia by the different periods of the COVID-19 pandemic. Incidence of primary care-coded anxiety disorders among individuals aged 18 to 64 years showed a sudden peak compared to expected rates in March, 2020, and was followed by reduction of diagnoses during the next months. Contrary to anxiety disorders, the incidence of depressive disorders sharply decreased among all sociodemographic groups reaching its minimum value in April, 2020. Based on observed data, incidence of anxiety and depressive disorders seemed to reach pre-lockdown levels in the first quarter of 2021. According to our model, which captures the underlying upward trend of diagnoses over time, the incidence of anxiety and depressive disorders did not fully reach what would have been expected. These results suggest that the effects of the measures implemented to control the spread of the pandemic are still negatively affecting the diagnoses of these mental health conditions in primary care.

Although the negative impact on the incidence of anxiety and depressive disorders appeared to occur across all ages, sex and SES, this was mostly pronounced among young adults or individuals living in the most deprived areas. Greater reductions were also found among females except for incident diagnoses of anxiety disorders during the post-lockdown period. These findings are particularly concerning, as these groups have been previously identified as experiencing worse mental health burden due to the pandemic.^4^ Incidence rate ratios provided by our study suggest that individuals aged 18 to 34 years were the only age group with significant increases in cases of anxiety and depressive disorders during the first trimester of 2021 compared to the pre-lockdown period. A possible explanation could be the increased impact of the pandemic on daily life, which has been heavily disrupted among the youth. Yet, they experienced the highest percentage of reductions according to our model during and after the lockdown period.

Reductions in common mental health diagnoses might be explained by the disruption of the normal functioning of healthcare systems due to the COVID-19 pandemic. In Catalonia, non-essential health activities were interrupted during lockdown in order to prioritize COVID-19 services, and health authorities advised against going to health centres except in the event of serious illness or urgent situations. While it is certainly true that healthcare systems reached capacity constraints, continuous dissemination of mass media messages reporting the strain of healthcare systems might have influenced health-care seeking behaviour. People might have delayed seeking care in order to avoid overburdening health systems or out of fear of contracting the disease.^23^ In such an extraordinary situation, low perceived legitimacy of mental issues might have also prevented those psychologically suffering to discern their emotional distress as an appropriate reason to seek care. In this line, we believe that a large number of people experiencing psychological distress might have developed coping strategies without seeking help in the public healthcare system. Prior evidence suggests that clinicians perceive the emotional distress of their patients, but have difficulties in specifying standardized psychiatric diagnoses.^17,24^ Consultations with a recorded diagnosis of a psychological problem take longer,^25^ and might require re-assessment of suspected cases to increase accuracy.^26^ It seems likely that the diagnoses detected in primary care during the pandemic might account for more severe cases of mental health issues. In addition, it is important to emphasize the increasing use of telemedicine in Catalonia, where the share of telemedicine-based visits has increased to 56.2%, from 15.4% in the pre-pandemic year.^28^ Since the pandemic, telemedicine tools serve a wider and more representative cross-section of the population in Catalonia, including underrepresented groups such as lower income individuals and those living in rural areas.^27^ While telemedicine has ensured the continuity of care of many health care processes during lockdown, it might face limitations generating diagnoses that require more assessments.^28^

A recent study performed in Central Catalonia (a health administrative region of Catalonia) found an average decline of 31.1% in new diagnoses compared to 2019.^28^ This study found an association between the intensity of the pandemic and the increased monthly declines in new diagnoses across ICD-10 groups, highlighting the strain on capacity of the healthcare system to identify and address the healthcare needs of the population during the pandemic. Another study performed in Catalonia has found a reduction in primary care visits associated with diagnoses related to chronic pathologies, while visits associated with diagnoses related to socio-economic and housing problems have increased.^29^ Both studies found moderate reductions in diagnoses of mental health conditions, which were lower than the ones observed for other diagnoses groups.^28,29^ Similar studies performed in the UK have used the same modelling approach to forecast incidence of anxiety and depressive disorders during lockdown and the first months of its aftermath.^22,30^ Larger reductions were also found for adults aged 18 to 64 years and for patients registered at practices in more deprived areas.^30^ In contrast to our findings, none of these studies have found an increase in new cases of anxiety disorders during lockdown. Comparison between studies performed in other countries render difficulties given the different strategies adopted to control de COVID-19 pandemic, which could have also played a role in the incidence of common mental disorders.

The reduced number of new diagnoses observed compared with the expected numbers obtained in this study are most likely to represent a large number of disorders that have gone undiagnosed and untreated. Previous studies suggest that diagnostic delays of anxiety and depressive disorders are associated with poorer outcomes.^12,13^ Possible consequences of this unmet need may include increased demand for mental health services, increased use of emergency departments for mental issues and heightened risk of suicide. The economic impacts of the pandemic will likely increase the risk of mental health problems and exacerbate health-care disparities.^31^ Monitoring patterns in primary care recording of common mental disorders, will provide crucial information to ensure health services can meet future demand. Data on psychotropic drug prescriptions, primary care contacts and secondary care referrals can provide valuable information on the management of anxiety and depressive disorders during the COVID-19 pandemic. Further research should also investigate the evolution of conditions over time using linkages to data from hospital emergency departments and outpatient mental health centers.

The main strengths of this study are the sample size and the real-world nature of the data. Our study provided data from a broadly representative setting, and included more than 4 million people registered in primary care in Catalonia. The data used were obtained from primary care electronic health records, which have been shown to be a useful tool for research in many areas, including COVID-19.^32,33^ In order to strengthen our results, we validated our modelling approach forecasting expected diagnoses from one year prior to the pandemic, obtaining consistent predictions compared to observed figures. Moreover, our study investigates the COVID-19 related effects up to March, 2021, providing data up to a year after the onset of the lockdown measures in Catalonia.

We acknowledge several limitations to our study. First, our data is limited to recorded diagnoses in primary care, which will have led to omit people with whom anxiety and depressive disorders were discussed or people who were treated for these conditions, but not entered as a diagnosis onto their clinical record. It is therefore likely that this is an underestimate of episodes of anxiety and depressive disorders. Second, reductions in diagnoses of anxiety and depressive disorders may have been underestimated as they were based on pre-pandemic data, and did not take into consideration an increase of incident cases of psychological distress due to the pandemic.

In conclusion, the sudden peak in primary-care recorded incident cases of anxiety disorders suggests increased mental distress at the onset of the lockdown measures in Catalonia. The marked reduction of incident diagnoses of anxiety and depressive disorders during the COVID-19 pandemic indicates untreated mental health problems given the increase in anxiety and depression observed in studies using self-reported data. Adults aged 18 to 34 and individuals living in most deprived areas might have greater levels of undetected need. Our findings may help to design public health interventions to target those particularly affected by the pandemic, as well as to prepare healthcare systems for greater demand for mental health services in the following months.

## Data Availability

In accordance with current European and national law, the data used in this study is only available for the researchers participating in this study. Thus, we are not allowed to distribute or make publicly available the data to other parties. However, researchers from public institutions can request data from SIDIAP if they comply with certain requirements. Further information is available online (https://www.sidiap.org/index.php/menu-solicitudesen/application-proccedure) or by contacting SIDIAP.

## Acknowledgment

The authors would like to acknowledge the efforts of all public health care workers in Catalonia during this global pandemic crisis. The authors would like to thank Daniel López-Codina and his team for assistance in fitting the model, and Miguel-Ángel Mayer for his contributions in the initial stages of the study.

## Contributors

BR, EB and TDS conceived and designed the study. SFB, MA, EB, and TDS mapped source data to the OMOP CDM. BR, EB and AP performed the statistical analysis. BR drafted the initial version of the manuscript. All authors (BR, AP, CR, SFB, MA, AB, CJA, LMP, EB, TDS) interpreted the results, critically reviewed the manuscript and approved the final version for submission.

## Ethics approval

This project was approved by the Clinical Research Ethics Committee of the IDIAPJGol (project code: 21/052-PCV).

## Transparency statement

The lead author affirms that the manuscript is an honest, accurate, and transparent account of the study being reported; that no important aspects of the study have been omitted; and that any discrepancies from the study as originally planned have been explained.

## Data availability statement

In accordance with current European and national law, the data used in this study is only available for the researchers participating in this study. Thus, we are not allowed to distribute or make publicly available the data to other parties. However, researchers from public institutions can request data from SIDIAP if they comply with certain requirements. Further information is available online (https://www.sidiap.org/index.php/menusolicitudesen/application-proccedure) or by contacting SIDIAP.

## Supplementary materials

**Supplementary Table 1.**
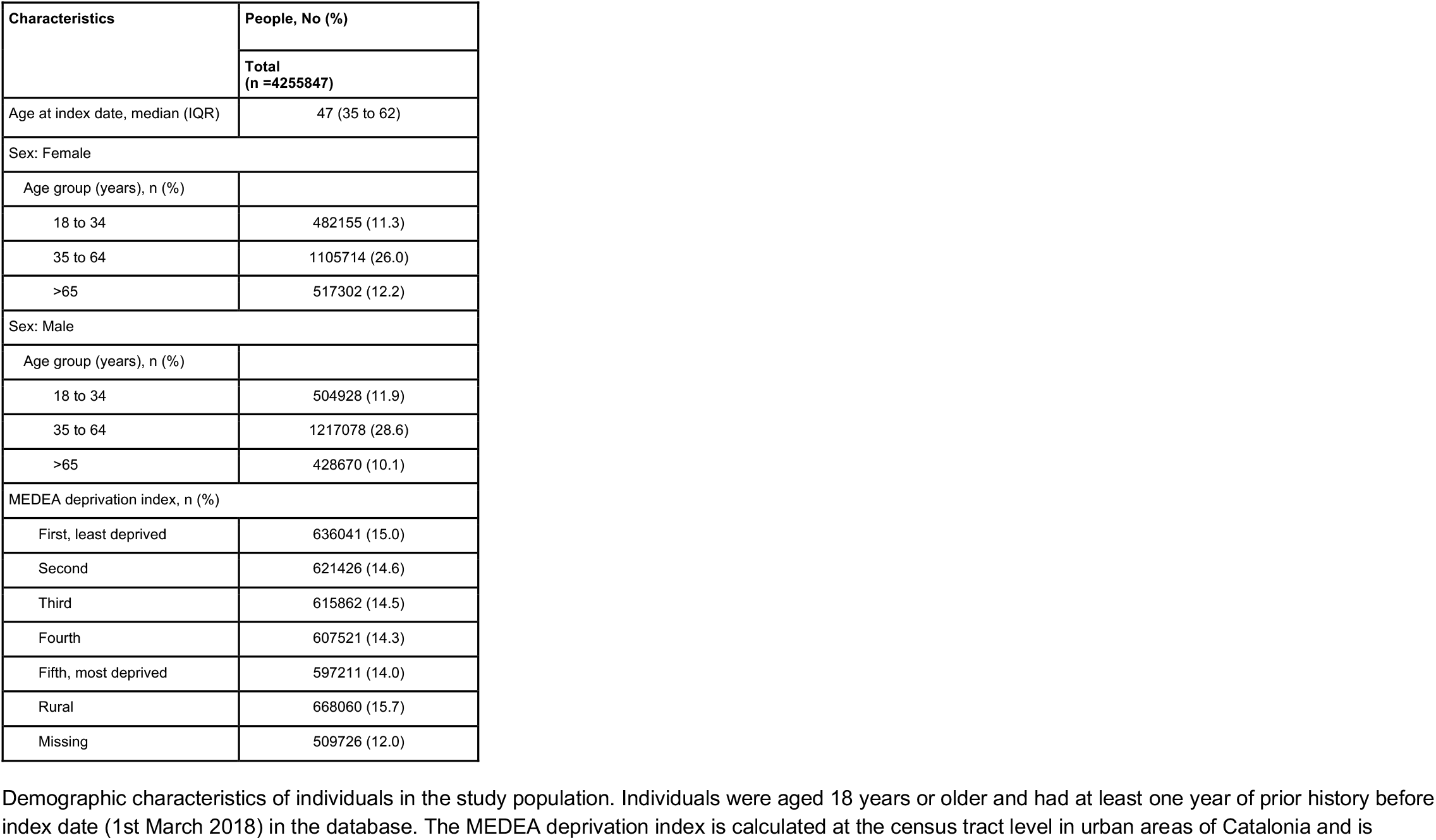
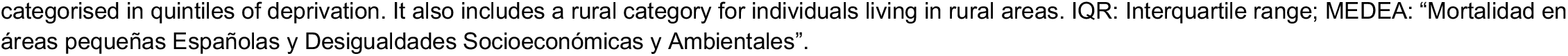
Characteristics of study participants

| Characteristics | People, No (%) |
| --- | --- |
|  | Total<br>(n =4255847) |
| Age at index date, median (IQR) | 47 (35 to 62) |
| Sex: Female |  |
| Age group (years), n (%) |  |
| 18 to 34 | 482155 (11.3) |
| 35 to 64 | 1105714 (26.0) |
| >65 | 517302 (12.2) |
| Sex: Male |  |
| Age group (years), n (%) |  |
| 18 to 34 | 504928 (11.9) |
| 35 to 64 | 1217078 (28.6) |
| >65 | 428670 (10.1) |
| MEDEA deprivation index, n (%) |  |
| First, least deprived | 636041 (15.0) |
| Second | 621426 (14.6) |
| Third | 615862 (14.5) |
| Fourth | 607521 (14.3) |
| Fifth, most deprived | 597211 (14.0) |
| Rural | 668060 (15.7) |
| Missing | 509726 (12.0) |

Demographic characteristics of individuals in the study population. Individuals were aged 18 years or older and had at least one year of prior history before index date (1st March 2018) in the database. The MEDEA deprivation index is calculated at the census tract level in urban areas of Catalonia and is
categorised in quintiles of deprivation. It also includes a rural category for individuals living in rural areas. IQR: Interquartile range; MEDEA: “Mortalidad en áreas pequeñas Españolas y Desigualdades Socioeconómicas y Ambientales”.

**Supplementary Table 2.**
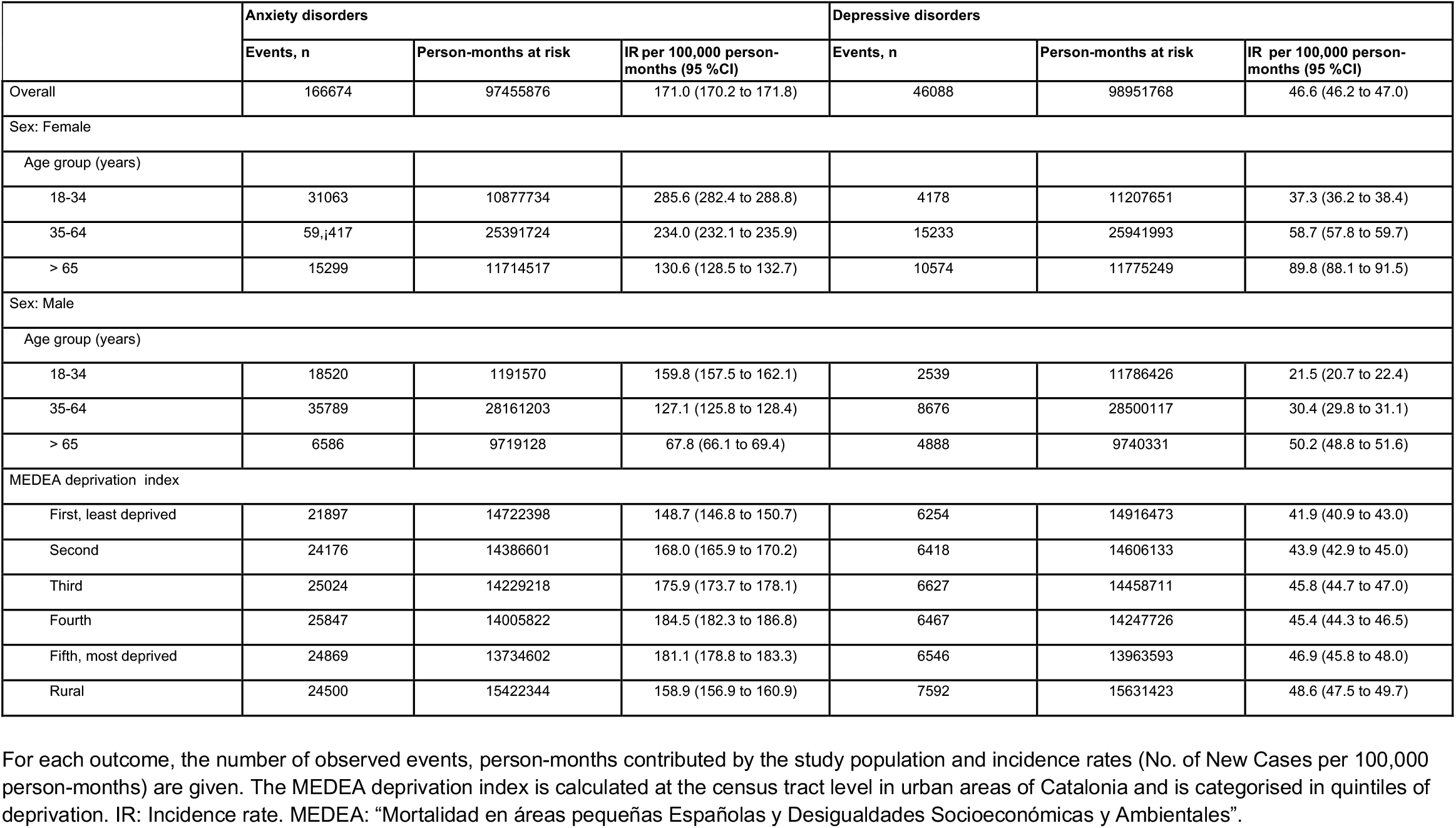
Incidence rates of anxiety and depressive disorders during the pre-lockdown period (March, 2018 - February, 2020)

|  | Anxiety disorders |  |  | Depressive disorders |  |  |
| --- | --- | --- | --- | --- | --- | --- |
|  | Events, n | Person-months at risk | IR per 100,000 person-months (95 %CI) | Events, n | Person-months at risk | IR per 100,000 person-months (95 %CI) |
| Overall | 166674 | 97455876 | 171.0 (170.2 to 171.8) | 46088 | 98951768 | 46.6 (46.2 to 47.0) |
| Sex: Female |  |  |  |  |  |  |
| Age group (years) |  |  |  |  |  |  |
| 18-34 | 31063 | 10877734 | 285.6 (282.4 to 288.8) | 4178 | 11207651 | 37.3 (36.2 to 38.4) |
| 35-64 | 59,417 | 25391724 | 234.0 (232.1 to 235.9) | 15233 | 25941993 | 58.7 (57.8 to 59.7) |
| > 65 | 15299 | 11714517 | 130.6 (128.5 to 132.7) | 10574 | 11775249 | 89.8 (88.1 to 91.5) |
| Sex: Male |  |  |  |  |  |  |
| Age group (years) |  |  |  |  |  |  |
| 18-34 | 18520 | 1191570 | 159.8 (157.5 to 162.1) | 2539 | 11786426 | 21.5 (20.7 to 22.4) |
| 35-64 | 35789 | 28161203 | 127.1 (125.8 to 128.4) | 8676 | 28500117 | 30.4 (29.8 to 31.1) |
| > 65 | 6586 | 9719128 | 67.8 (66.1 to 69.4) | 4888 | 9740331 | 50.2 (48.8 to 51.6) |
| MEDEA deprivation index |  |  |  |  |  |  |
| First, least deprived | 21897 | 14722398 | 148.7 (146.8 to 150.7) | 6254 | 14916473 | 41.9 (40.9 to 43.0) |
| Second | 24176 | 14386601 | 168.0 (165.9 to 170.2) | 6418 | 14606133 | 43.9 (42.9 to 45.0) |
| Third | 25024 | 14229218 | 175.9 (173.7 to 178.1) | 6627 | 14458711 | 45.8 (44.7 to 47.0) |
| Fourth | 25847 | 14005822 | 184.5 (182.3 to 186.8) | 6467 | 14247726 | 45.4 (44.3 to 46.5) |
| Fifth, most deprived | 24869 | 13734602 | 181.1 (178.8 to 183.3) | 6546 | 13963593 | 46.9 (45.8 to 48.0) |
| Rural | 24500 | 15422344 | 158.9 (156.9 to 160.9) | 7592 | 15631423 | 48.6 (47.5 to 49.7) |

For each outcome, the number of observed events, person-months contributed by the study population and incidence rates (No. of New Cases per 100,000 person-months) are given. The MEDEA deprivation index is calculated at the census tract level in urban areas of Catalonia and is categorised in quintiles of deprivation. IR: Incidence rate. MEDEA: “Mortalidad en áreas pequeñas Españolas y Desigualdades Socioeconómicas y Ambientales”.

**Supplementary Table 3.** Incidence rates of anxiety and depressive disorders during the lockdown period (March, 2020 - June, 2020)

|  | Anxiety disorders |  |  | Depressive disorders |  |  |
| --- | --- | --- | --- | --- | --- | --- |
|  | Events, n | Person-months at risk | IR per 100,000 person-months (95 %CI) | Events, n | Person-months at risk | IR per 100,000 person-months (95 %CI) |
| Overall | 30982 | 153.655.78 | 201.6 (199.4 to 203.9) | 4691 | 159.179.02 | 29.5 (28.6 to 30.3) |
| Sex: Females |  |  |  |  |  |  |
| Age group (years) |  |  |  |  |  |  |
| 18-34 | 4808 | 1685994 | 285.2 (277.2 to 293.3) | 367 | 1805057 | 20.3 (18.3 to 5) |
| 35-64 | 11787 | 4017488 | 293.4 (288.1 to 298.7) | 1620 | 4222691 | 38.4 (36.5 to 40.3) |
| > 65 | 2735 | 1816718 | 150.5 (145.0 to 156.3) | 1031 | 183.9817 | 56.0 (52.7 to 59.6) |
| Sex: Males |  |  |  |  |  |  |
| Age group (years) |  |  |  |  |  |  |
| 18-34 | 3113 | 1834608 | 169.7(163.8 to 175.7) | 234 | 1906042 | 12.3 (10.8 to 14.0) |
| 35-64 | 7137 | 4505518 | 158.4 (154.8 to 162.1) | 912 | 4630125 | 19.7 (18.4 to 21.0) |
| > 65 | 1402 | 1505252 | 93.1 (88.3 to 98.1) | 527 | 1514170 | 34.8 (31.9 to 37.9) |
| MEDEA deprivation index |  |  |  |  |  |  |
| First, least deprived | 4049 | 2352652 | 172.1 (166.8 to 177.5) | 671 | 2424658 | 27.7 (25.6 to 29.8) |
| Second | 4596 | 2296889 | 200.1 (194.4 to 206.0) | 669 | 2379030 | 28.1 (26.0 to 30.3) |
| Third | 4806 | 2267702 | 211.9 (206.0 to 218.0) | 655 | 2352638 | 27.8 (25.7 to 30.1) |
| Fourth | 4908 | 2226456 | 220.4 (214.3 to 226.7) | 639 | 2315857 | 27.6 (25.5 to 29.8) |
| Fifth, most deprived | 4685 | 2177422 | 215.2 (209.0 to 221.4) | 593 | 2262426 | 26.2 (24.1 to 28.4) |
| Rural | 4642 | 2454530 | 189.1 (183.7 to 194.6) | 847 | 2532640 | 33.4 (31.2 to 35.8) |

For each outcome, the number of observed events, person-months contributed by the study population and incidence rates (No. of New Cases per 100,000 person-months) are given. The MEDEA deprivation index is calculated at the census tract level in urban areas of Catalonia and is categorised in quintiles of deprivation. IR: Incidence rate. MEDEA: “Mortalidad en áreas pequeñas Españolas y Desigualdades Socioeconómicas y Ambientales”.

**Supplementary Table 4.**
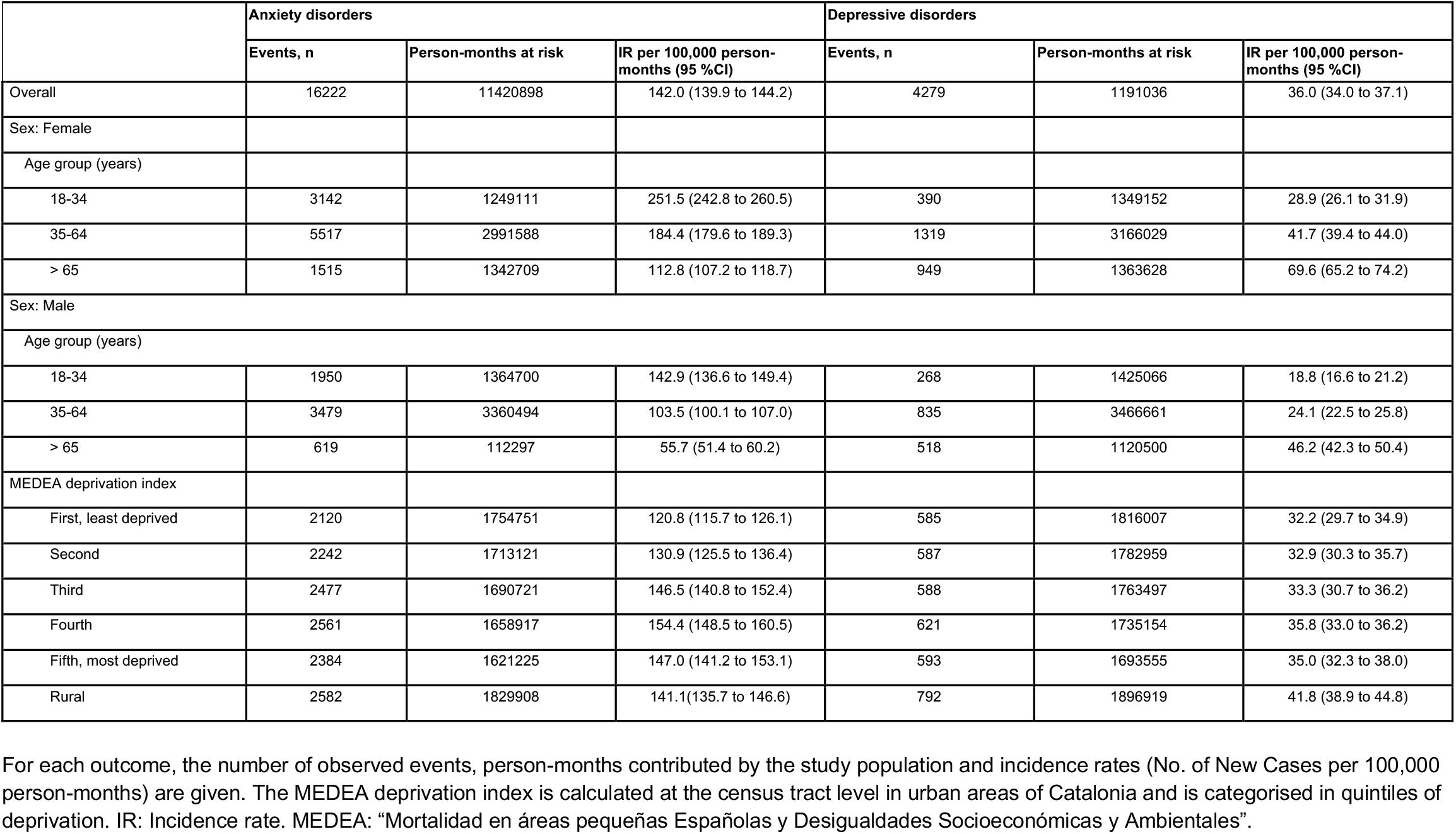
Incidence rates of anxiety and depressive disorders during the first trimester of the post-lockdown period (July, 2020 - September, 2020)

|  | Anxiety disorders |  |  | Depressive disorders |  |  |
| --- | --- | --- | --- | --- | --- | --- |
|  | Events, n | Person-months at risk | IR per 100,000 person-months (95 %CI) | Events, n | Person-months at risk | IR per 100,000 person-months (95 %CI) |
| Overall | 16222 | 11420898 | 142.0 (139.9 to 144.2) | 4279 | 1191036 | 36.0 (34.0 to 37.1) |
| Sex: Female |  |  |  |  |  |  |
| Age group (years) |  |  |  |  |  |  |
| 18-34 | 3142 | 1249111 | 251.5 (242.8 to 260.5) | 390 | 1349152 | 28.9 (26.1 to 31.9) |
| 35-64 | 5517 | 2991588 | 184.4 (179.6 to 189.3) | 1319 | 3166029 | 41.7 (39.4 to 44.0) |
| > 65 | 1515 | 1342709 | 112.8 (107.2 to 118.7) | 949 | 1363628 | 69.6 (65.2 to 74.2) |
| Sex: Male |  |  |  |  |  |  |
| Age group (years) |  |  |  |  |  |  |
| 18-34 | 1950 | 1364700 | 142.9 (136.6 to 149.4) | 268 | 1425066 | 18.8 (16.6 to 21.2) |
| 35-64 | 3479 | 3360494 | 103.5 (100.1 to 107.0) | 835 | 3466661 | 24.1 (22.5 to 25.8) |
| > 65 | 619 | 112297 | 55.7 (51.4 to 60.2) | 518 | 1120500 | 46.2 (42.3 to 50.4) |
| MEDEA deprivation index |  |  |  |  |  |  |
| First, least deprived | 2120 | 1754751 | 120.8 (115.7 to 126.1) | 585 | 1816007 | 32.2 (29.7 to 34.9) |
| Second | 2242 | 1713121 | 130.9 (125.5 to 136.4) | 587 | 1782959 | 32.9 (30.3 to 35.7) |
| Third | 2477 | 1690721 | 146.5 (140.8 to 152.4) | 588 | 1763497 | 33.3 (30.7 to 36.2) |
| Fourth | 2561 | 1658917 | 154.4 (148.5 to 160.5) | 621 | 1735154 | 35.8 (33.0 to 36.2) |
| Fifth, most deprived | 2384 | 1621225 | 147.0 (141.2 to 153.1) | 593 | 1693555 | 35.0 (32.3 to 38.0) |
| Rural | 2582 | 1829908 | 141.1(135.7 to 146.6) | 792 | 1896919 | 41.8 (38.9 to 44.8) |

For each outcome, the number of observed events, person-months contributed by the study population and incidence rates (No. of New Cases per 100,000 person-months) are given. The MEDEA deprivation index is calculated at the census tract level in urban areas of Catalonia and is categorised in quintiles of deprivation. IR: Incidence rate. MEDEA: “Mortalidad en áreas pequeñas Españolas y Desigualdades Socioeconómicas y Ambientales”.

**Supplementary Table 5.** Incidence rates of anxiety and depressive disorders during the second trimester of the post-lockdown period (October, 2020 - December, 2020)

|  | Anxiety disorders |  |  | Depressive disorders |  |  |
| --- | --- | --- | --- | --- | --- | --- |
|  | Events, n | Person-months at risk | IR per 100,000 person-months (95 %CI) | Events, n | Person-months at risk | IR per 100,000 person-months (95 %CI) |
| Overall | 16873 | 112.939.31 | 149.4 (147.2 to 151.7) | 4728 | 11800273 | 40.1 (38.9 to 41.2) |
| Sex: Female |  |  |  |  |  |  |
| Age group (years) |  |  |  |  |  |  |
| 18-34 | 3296 | 1231819 | 267.6 (258.5 to 276.9) | 514 | 1340217 | 38.4 (35.1 to 41.8) |
| 35-64 | 6143 | 2962125 | 207.4 (202.2 to 212.6) | 1610 | 3150017 | 51.1 (48.6 to 53.7) |
| > 65 | 1600 | 1320435 | 121.2 (115.3 to 127.3) | 992 | 1343120 | 73.9 (69.3 to 78.6) |
| Sex: Male |  |  |  |  |  |  |
| Age group (years) |  |  |  |  |  |  |
| 18-34 | 1774 | 1351418 | 131.3 (125.2 to 137.5) | 277 | 1416371 | 19.6 (17.3 to 22.0) |
| 35-64 | 3455 | 3334606 | 103.6 (100.2 to 107.1) | 867 | 3448456 | 25.1 (23.5 to 26.9) |
| > 65 | 605 | 1093528 | 55.3 (51.0 to 59.9) | 468 | 1102093 | 42.5 (38.7 to 46.5) |
| MEDEA deprivation index |  |  |  |  |  |  |
| First, least deprived | 2357 | 1741170 | 135.4 (130.0 to 140.9) | 642 | 1807488 | 35.5 (32.8 to 38.4) |
| Second | 2541 | 1700154 | 149.5 (143.7 to 155.4) | 685 | 1775487 | 38.6 (35.7to 41.6) |
| Third | 2531 | 1677402 | 150.9 (153.3 to 165.5) | 686 | 1755919 | 39.1 (36.2 to 42.1) |
| Fourth | 2621 | 1645206 | 159.3 (153.3 to 165.5) | 692 | 1727280 | 40.1 (37.1 to 43.2) |
| Fifth, most deprived | 2400 | 1607406 | 149.3 (143.4 to 155.4) | 669 | 1685144 | 39.7 (36.7 to 42.8) |
| Rural | 2620 | 1813723 | 144.5 (139.0 to 150.1) | 789 | 1886237 | 41.8 (39.0to 44.9) |

For each outcome, the number of observed events, person-months contributed by the study population and incidence rates (No. of New Cases per 100,000 person-months) are given. The MEDEA deprivation index is calculated at the census tract level in urban areas of Catalonia and is categorised in quintiles of deprivation. IR: Incidence rate. MEDEA: “Mortalidad en áreas pequeñas Españolas y Desigualdades Socioeconómicas y Ambientales”.

**Supplementary Table 6.** Incidence rates of anxiety and depressive disorders during the third trimester of the post-lockdown period (January, 2021 - March, 2021)

|  | Anxiety disorders |  |  | Depressive disorders |  |  |
| --- | --- | --- | --- | --- | --- | --- |
|  | Events, n | Person-months at risk | IR per 100,000 person-months (95 %CI) | Events, n | Person-months at risk | IR per 100,000 person-months (95 %CI) |
| Overall | 19171 | 10886624 | 176.1 (173.6 to 178.6) | 5319 | 11417692 | 46.6 (45.3 to 47.9) |
| Gender: Female |  |  |  |  |  |  |
| Age group (years) |  |  |  |  |  |  |
| 18-34 | 3815 | 118.255.798.291.721 | 322.6 (312.4 to 333.0) | 556 | 1296588 | 42.9 (39.4 to 46.6) |
| 35-64 | 6773 | 286.245.321.944.809 | 236.6 (231.0 to 242.3) | 1812 | 3059473 | 59.2 (56.5 to 62.0) |
| > 65 | 1635 | 126.584.747.043.364 | 129.2 (123.0 to 135.6) | 1044 | 1289780 | 80.9 (76.1 to 86.0) |
| Gender: Male |  |  |  |  |  |  |
| Age group (years) |  |  |  |  |  |  |
| 18-34 | 2174 | 1302697 | 166.9 (159.9 to 174.1) | 349 | 1370706 | 25.5 (22.9 to 28.3) |
| 35-64 | 4132 | 3226223 | 128.1 (124.2 to 132.0) | 1081 | 3345388 | 32.3 (30.4 to 34.3) |
| > 65 | 642 | 1046845 | 61.3 (56.7 to 66.3) | 477 | 1055756 | 45.2 (41.2 to 49.4) |
| Deprivation |  |  |  |  |  |  |
| First, least deprived | 2560 | 1685241 | 151.9 (146.1 to 157.9) | 737 | 1755143 | 42.0 (39.0 to 45.1) |
| Second | 2921 | 1646843 | 177.4 (171.0 to 183.9) | 780 | 1726091 | 45.2 (42.1 to 48.5) |
| Third | 2883 | 1624045 | 177.5 (171.1 to 184.1) | 756 | 1706526 | 44.3 (41.2 to 47.6) |
| Fourth | 2934 | 1591422 | 184.4 (177.8 to 191.2) | 788 | 1677497 | 47.0 (43.8 to 50.4) |
| Fifth, most deprived | 2848 | 1552548 | 183.4 (176.8 to 190.3) | 779 | 16.34047 | 47.7 (44.4 to 51.1) |
| Rural | 2984 | 1755264 | 170.0 (164.0 to 176.2) | 930 | 1831618 | 50.8 (47.6 to 54.1) |

For each outcome, the number of observed events, person-months contributed by the study population and incidence rates (No. of New Cases per 100,000 person-months) are given. The MEDEA deprivation index is calculated at the census tract level in urban areas of Catalonia and is categorised in quintiles of deprivation. IR: Incidence rate. MEDEA: “Mortalidad en áreas pequeñas Españolas y Desigualdades Socioeconómicas y Ambientales”.

**Supplementary Figure 1.**
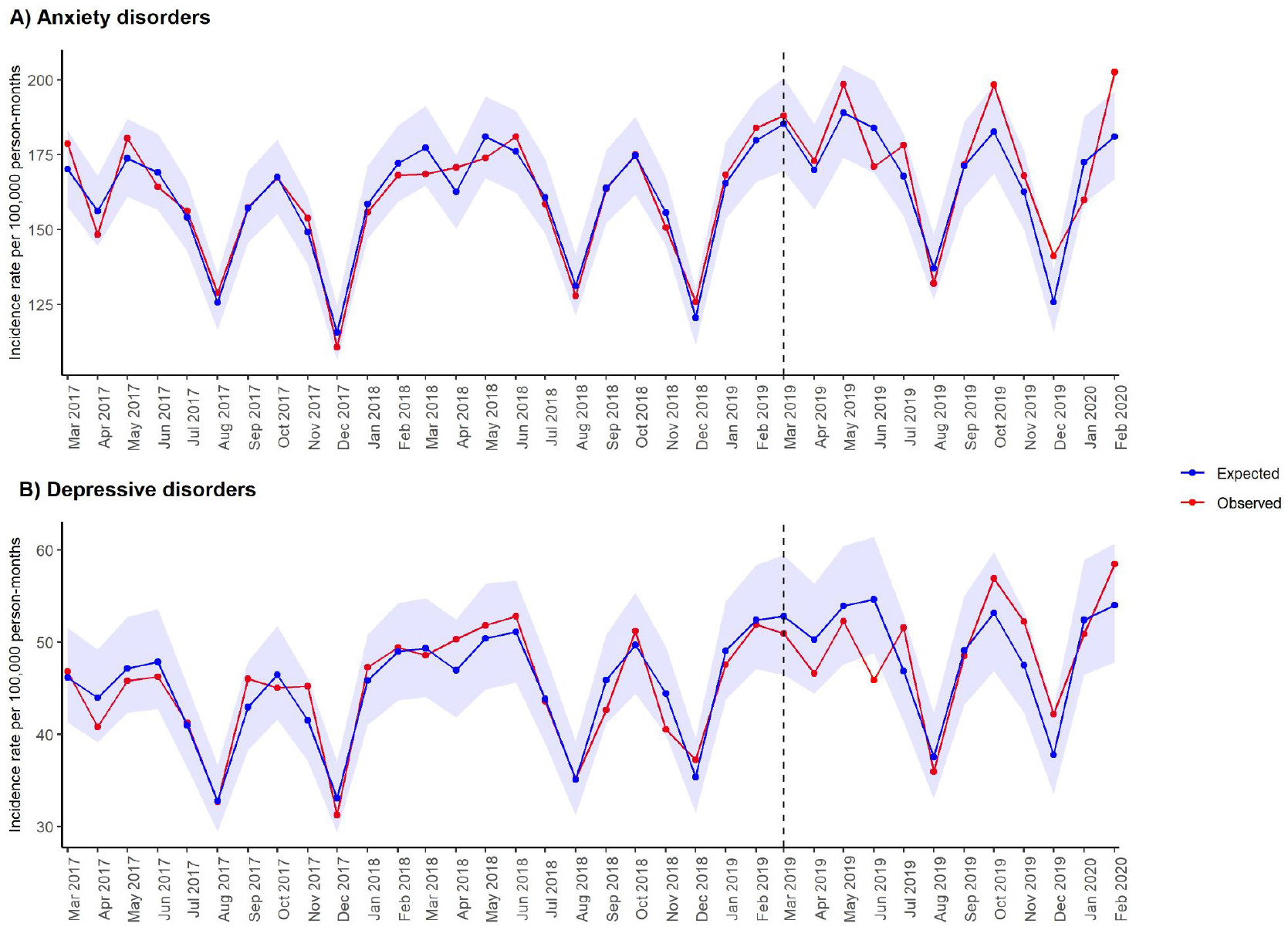
Expected and observed incidence of anxiety and depressive disorders in primary care in Catalonia (March, 2017– February, 2020). Validation of the modelling approach for overall incidence rates of anxiety and depressive disorders. Number of expected cases (95% PI) were estimated with negative binomial models, using data from 1 March, 2017 to 28 February, 2019. Shaded areas in blue represent 95 % PI. Vertical lines show 1 March, 2019.

**Supplementary Figure 2.**
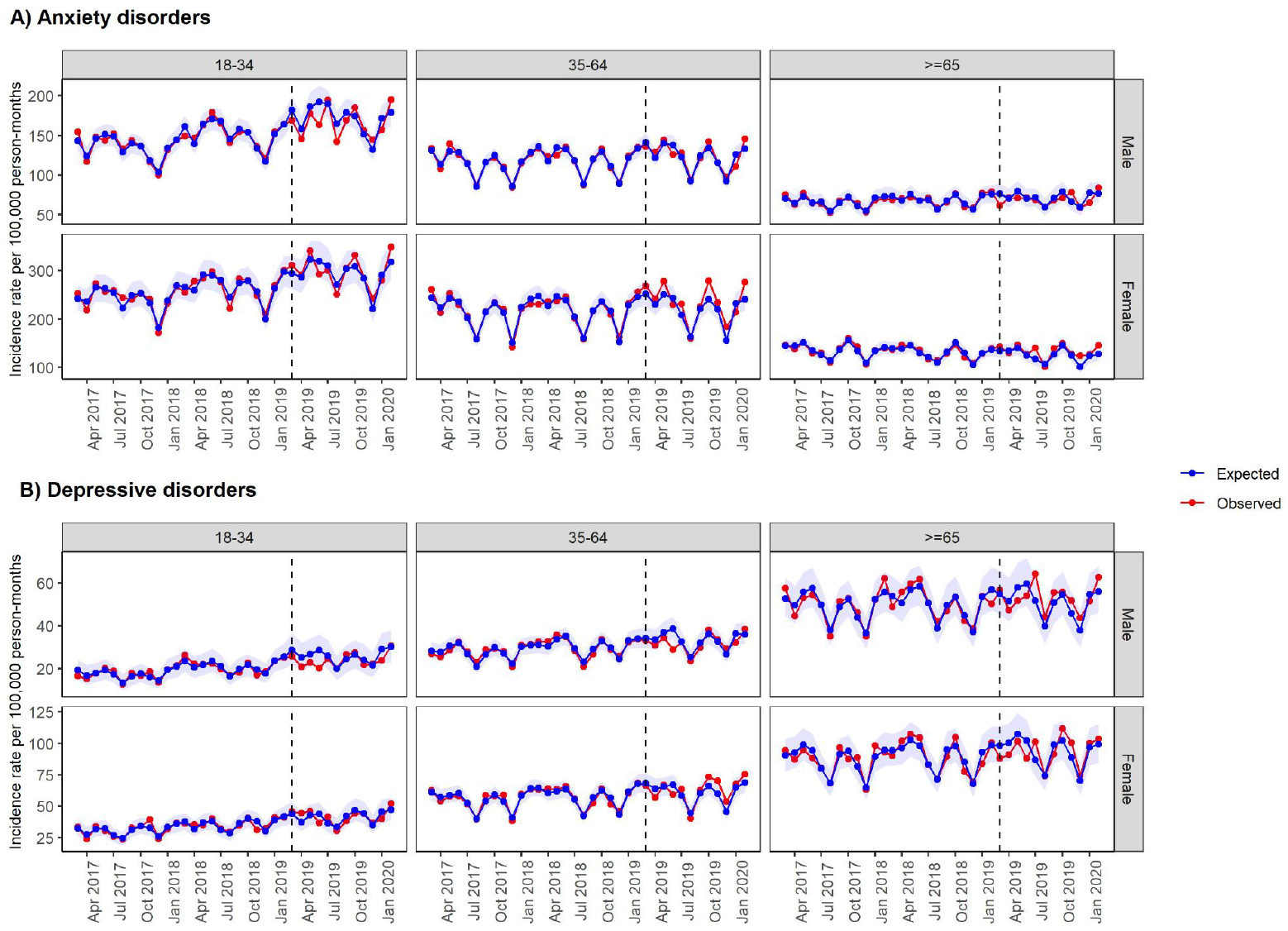
Expected and observed incidence of anxiety and depressive disorders stratified by age group and gender in primary care in Catalonia (March, 2017– February, 2020). Validation of the modelling approach for incidence rates of anxiety and depressive disorders stratified by age and gender. Number of expected cases (95% PI) were estimated with negative binomial models, using data from 1 March, 2017 to 28 February, 2019. Vertical lines show 1 March, 2019. Shaded areas in blue represent 95 % PI.

**Supplementary Figure 3.**
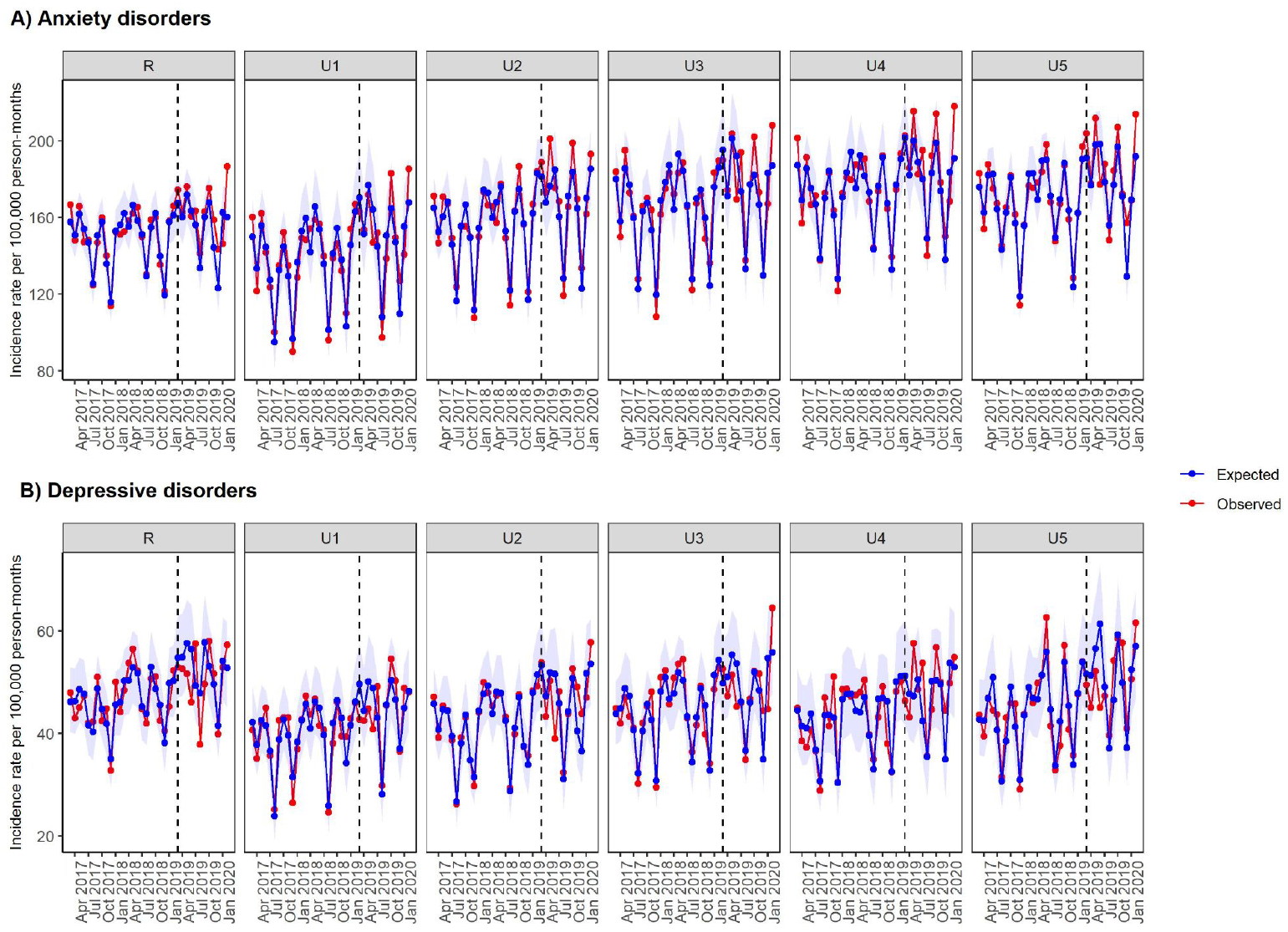
Expected and observed incidence of anxiety and depressive disorders stratified by MEDEA deprivation index in primary care in Catalonia (March, 2017– February, 2020). Validation of the modelling approach for incidence rates of anxiety and depressive disorders stratified by age and gender. Number of expected cases (95% PI) were estimated with negative binomial models, using data from 1 March, 2017 to 28 February, 2019. Shaded areas in blue represent 95 % PI. Vertical lines show 1 March, 2019. The MEDEA deprivation index is calculated at the census tract level in urban areas of Catalonia and is categorised in quintiles of deprivation. It also includes a rural category for individuals living in rural areas.

